# Genome-wide polygenic score with *APOL1* risk genotypes predicts chronic kidney disease across major continental ancestries

**DOI:** 10.1101/2021.10.25.21265398

**Authors:** Atlas Khan, Michael C. Turchin, Amit Patki, Vinodh Srinivasasainagendra, Ning Shang, Rajiv Nadukuru, Alana C. Jones, Edyta Malolepsza, Ozan Dikilitas, Iftikhar J. Kullo, Daniel J. Schaid, Elizabeth Karlson, Tian Ge, James B. Meigs, Jordan W. Smoller, Christoph Lange, David R. Crosslin, Gail Jarvik, Pavan Bhatraju, Jacklyn N. Hellwege, Paulette Chandler, Laura Rasmussen Torvik, Alex Fedotov, Cong Liu, Christopher Kachulis, Noura S. Abul-Husn, Judy H. Cho, Iuliana Ionita-Laza, Ali G. Gharavi, Wendy K. Chung, George Hripcsak, Chunhua Weng, Girish Nadkarni, Marguerite R. Irvin, Hemant K. Tiwari, Eimear E. Kenny, Nita A. Limdi, Krzysztof Kiryluk

## Abstract

**Introduction:** Chronic kidney disease (CKD) is a common complex condition associated with significant morbidity and mortality in the US and worldwide. Early detection is critical for effective prevention of kidney disease progression. Polygenic prediction of CKD could enhance screening and prevention of kidney disease progression, but this approach has not been optimized for risk prediction in ancestrally diverse populations.

**Methods:** We developed and validated a genome-wide polygenic score (GPS) for CKD defined by estimated glomerular filtration rate (eGFR) <60 mL/min/1.73m^2^ using common variant association statistics from GWAS for eGFR combined with information on *APOL1* risk genotypes. The score was designed to ensure transferability across major continental ancestries, genotyping platforms, imputation panels, and phenotyping strategies, and was tested following ClinGen guidelines. The polygenic component of the score was developed and optimized using 28,047 cases and 251,772 controls (70% of UK Biobank participants of European ancestry), while the weights for *APOL1* effects were derived based on UK Biobank participants of African ancestry (967 cases and 6,191 controls). We tested the performance of the score in 15 independent testing cohorts, including 3 cohorts of European ancestry (total 23,364 cases and 117,883 controls), 6 cohorts of African ancestry (4,268 cases and 10,276 controls), 4 cohorts of Asian ancestry (1,030 cases and 9,896 controls), and 2 Hispanic/Latinx cohorts (1,492 cases and 2,984 controls).

**Results:** We demonstrated the risk score transferability with reproducible performance across all independent testing cohorts. In the meta-analyses, disease odds ratios per standard deviation of the score were estimated at 1.49 (95%CI: 1.47-1.50, P<1.0E-300) for European, 1.32 (95%CI: 1.26-1.38, P=1.8E-33) for African, 1.59 (95%CI: 1.52-1.67, P=1.3E-30) for Asian, and 1.42 (95%CI: 1.33-1.51, P=4.1E-14) for Latinx cohorts. The top 2% cutoff of the GPS was associated with nearly 3-fold increased risk of CKD across all major ancestral groups, the degree of risk that is equivalent to a positive family history of kidney disease. In African-ancestry cohorts, *APOL1* risk genotype and the polygenic risk components of the GPS had additive effects on the risk of CKD with no significant interactions. We also observed that individuals of African ancestry had a significantly higher polygenic risk score for CKD compared to other populations, even without accounting for *APOL1* variants.

**Conclusions:** By combining *APOL1* risk genotypes with the available GWAS for renal function, we designed, optimized, and validated a GPS predictive of CKD across four major continental ancestries. With the upper tail of the GPS distribution associated with disease risk equivalent to a positive family history, this score could be used for clinically meaningful risk stratification.

## INTRODUCTION

Chronic kidney disease (CKD) affects 10-16% of general population and has high morbidity and mortality^1-3^. In the US, CKD disproportionally affects African Americans (16.3%) when compared to European Americans (12.7%), Asian Americans (12.9%), or Hispanic Americans (13.6%) (https://www.cdc.gov/kidneydisease/publications-resources/ckd-national-facts.html). CKD stage 3 or greater is defined by a chronic loss of glomerular filtration rate (GFR) to under 60 mL/min/1.73m^2^. Because this definition is based on estimated kidney function rather than markers of specific kidney injury, it captures an etiologically heterogeneous set of primary and secondary kidney disorders. As expected for a highly heterogeneous trait, CKD has a complex determination with both genetic and environmental contributions. The observational heritability of CKD based on large-scale analysis of medical records ranged from 25-44% depending on self-reported race and ethnicity, with higher heritability estimated for pedigrees with majority of members self-identifying as Black^4^. These heritability estimates are generally consistent with prior smaller family-based studies of CKD and glomerular filtration rate^5-11^.

The substantial heritability of CKD is attributed to both monogenic^12-14^ and polygenic causes^15-19^. Moreover, in individuals of African ancestry, two common risk alleles (G1 and G2) in *Apolipoprotein L1* (*APOL1*) gene have been described to convey a large effect on the risk of kidney disease^20,21^. While heterozygotes for G1 or G2 alleles appear to be protected from trypanosomal sleeping sickness, kidney disease risk is conveyed under a recessive model in carriers of two risk alleles (G1G1, G2G2, or G1G2). Because of the selective pressure exerted by endemic trypanosomal species in certain parts of eastern and western Africa, G1 and G2 alleles are observed almost exclusively individuals whose ancestry can be linked to those areas^22,23^. In the US population, frequency of *APOL1* risk genotypes is estimated at approximately 15% in African Americans, 0.5-2% in Hispanic Americans, and <0.01% in European Americans^24^. These differences may be contributing to the higher prevalence of CKD in African Americans in the US, but additional non-*APOL1* genetic risk factors have not yet been elucidated.

Genome-wide polygenic scores (GPS) have emerged as promising tools for genetic risk stratification that can enhance traditional risk models for complex diseases. This approach has been applied to a variety of traits, including coronary artery disease^25-35^, type 2 diabetes^28-30^, hypertension^36,37^, obesity^38^, schizophrenia^33-35^, and malignancies including breast, colorectal, prostate, and lung cancers^39-46^. One of the major limitations of the GPS approach is that existing GWAS are based predominantly on European cohorts and, as a result, most GPS do not perform well in more diverse cohorts, or in individuals with admixed ancestry^47^. Similar to other complex traits, GWAS for kidney function involved predominantly European cohorts. The latest study involved 765,348 participants, 75% of which were European, 23% East Asian, 2% African American, and <1% Hispanic^17^. Notably, this study did not capture the effects of *APOL1* risk variants because of their recessive inheritance and very low frequencies in non-African populations.

The objective of the present study is to test if the existing knowledge on polygenic contributions to renal function is sufficient to build a clinical risk predictor for moderate-to-advanced CKD with adequate performance across diverse ancestral groups. We specifically aimed to design, optimize, and test a new GPS for clinical risk prediction of kidney disease that maximizes the performance across major continental ancestries. We combined information on *APOL1* risk genotypes with the latest GWAS for renal function to formulate a GPS that can reliably discriminate moderate to advanced CKD (stage 3 or greater) from population controls. In our approach, we took advantage of the power of the existing GWAS for a quantitative biomarker of renal function (serum creatinine-based eGFR) to predict a disease state. To demonstrate transferability across different genotyping and imputation platforms, and to document comparable predictive performance by ancestry, we performed rigorous testing of our GPS in 15 independent and ancestrally diverse case-control cohorts following ClinGen standards^48^.

## METHODS

### Study cohorts: genotyping, imputation, and quality control analyses

#### Electronic Medical Records and Genomics (eMERGE)

The eMERGE network provides access to EHR information linked to GWAS data for 102,138 individuals; detailed quality control analyses of genetic data have been described previously^4,49,50^. Briefly, GWAS datasets were imputed using the latest multiethnic Haplotype Reference Consortium (HRC) panel using Michigan Imputation Server^51^. The imputation was performed in 81 batches across the 12 contributing medical centers participating in eMERGE-I, II, and III. For post-imputation analyses, we included only markers with minor allele frequency (MAF)≥ 0.01 and *R*^2^ ≥ 0.8 in ≥ 75% of batches. A total of 7,529,684 variants were retained for the GPS analysis. For principal component analysis (PCA), we used FlashPCA^52^ on a set of 48,509 common (MAF>0.01) and independent variants (pruned in PLINK with --indep-pairwise 500 50 0.05 command). The G1 (1072A>G (rs73885319) and 1200T>G (rs60910145)) and G2 (1212-del6 (rs71785313)) alleles of *APOL1* were imputed separately using the TOPMed imputation server^53^. The allelic frequencies of G1 and G2 alleles were comparable to previous studies^54^ and were summarized in **Supplementary Table 1**. The analyses were performed using a combination of VCFtools, PLINK, and custom scripts in PYTHON and R^55-57^.

#### UK Biobank (UKBB)

UKBB is a large prospective cohort based in the United Kingdom that enrolled individuals ages 40-69 for the purpose of genetic studies^58^. This cohort is comprised of 488,377 individuals recruited since 2006, genotyped with high-density SNP arrays, and linked to electronic health record data. All individuals underwent genome-wide genotyping with UK Biobank Axiom array from Affymetrix and UK BiLEVE Axiom arrays (∼825,000 markers). Genotype imputation was carried out using a 1000 Genomes reference panel with IMPUTE4 software^59-61^. We then applied QC filters similar to eMERGE-III, retaining 9,233,643 common (MAF ≥ 0.01)variants imputed with high confidence (*R*^*^ ≥ 0.8). For principal component analysis by FlashPCA^52^, we used a set of 35,226 variants that were common (MAF>0.01) and pruned using the following command in PLINK --indep-pairwise 500 50 0.05. Similar to the eMERGE-III datasets, the *APOL1* G1 and G2 alleles were imputed separately using the TOPMed imputation server^53^.

#### Bio*Me* Biobank

The Bio*Me* Biobank is an electronic health record (EHR)-linked biorepository that has been enrolling participants non-selectively from across the Mount Sinai Health System (MSHS) in New York City since 2007. There are currently over 60,000 participants enrolled in Bio*Me* under and Institutional Review Board (IRB) approved study protocol (IRB 07–0529). Participants consent to provide DNA and plasma samples linked to their EHRs. Participants provide additional information on self-reported ancestry, personal and family health history through questionnaires administered upon enrollment. A total of 32,595 Bio*Me* participants were genotyped on the Illumina Global Screening Array (GSA) through a collaboration with Regeneron Genetics Center and 11,953 on the Illumina Global Diversity Array (GDA) through a collaboration with Sema4. Population groups were determined by self-reported race/ethnicity as published previosuly^62^. Participants were removed if genotype missing rates were > 5%, and if sex or ancestry information were missing. Variants were removed if genotype missing rates were >5% and if HWE was less than P < 1.00E-5 for the GSA data and p < 1.00E-6 for the GDA data within each ancestry group. Imputation (including G1 and G2 variants in *APOL1*) was performed using the TOPMed Imputation Server and the TOPMed Freeze 8 reference panel. Post-imputation variants with quality scores <0.7 were removed. Participants younger than 40 years old and individuals detected to be cryptically related (2nd degree or above) were removed from the analysis. We additionally removed a subset of Bio*Me* participants that were included in the original CKDGen GWAS discovery dataset^17^. After all QC steps, there were 9,154 Bio*Me* participants of European ancestry, 7,318 African ancestry, 11,606 Latinx ancestry, and 843 East Asian ancestry included in the analysis.

#### Reasons for Geographic and Racial Differences in Stroke Study (REGARDS)

REGARDS is a population-based, longitudinal study of incident stroke and associated risk factors of over 30,000 Black and White adults aged 45 years or older from all 48 contiguous US states and the District of Columbia^63^. REGARDS was designed to investigate reasons underlying the higher rate of stroke mortality among self-reported Black compared to White participants, as well as why residents of the Southeastern US had worse death rates compared to other US regions. By design, participants were oversampled if they were residents of the stroke belt or if they were Black^6363626161^. Participants completed a computer-assisted telephone interview to collect demographic information and medication adherence, and an in-home visit for blood pressure measurements and collection of blood and urine samples and have been contacted at six-month intervals to obtain information on incident stroke or secondary outcomes. Genotyping was performed on 8,916 self-identified Black participants using Illumina MEGA-EX arrays and imputed using the NHLBI TOPMed reference panel (Freeze 8). Participants were excluded with call rates less than 95%, if they were internal duplicates, had sex mismatches, or were outliers on principal component analysis (outside of 6 standard deviations), resulting in 8,669 participants with genotypes available for analysis. Imputed variants were inspected for their imputation quality scores (R^2^) and it was noted that more than 99% of the variants with MAF>1% had an imputation quality of 0.6 or higher. Given the high-quality of imputation for variants of MAF>1%, in order to retain maximum overlap of variants with the SNPs with PRS weights, genotypes with genotypic probability of 0.9 or higher were retained. *APOL1* alleles (G1 [rs73885319A>G, S342G] and G2 [rs71785313 TTATAA/– N388Y389/–]) were genotyped directly using TaqMan SNP Genotyping Assays (Applied Biosystems/ThermoFisher Scientific).

#### The Hypertension Genetic Epidemiology Network (HyperGEN)

HyperGEN is a cross-sectional, population-based study and component of the NHLBI Family Blood Pressure Program that was designed to identify genetic risk factors for hypertension and target end-organ damage due to hypertension^64^. The cohort is composed of White and Black sibships in which at least two siblings were diagnosed with hypertension (defined as either self-reported use of antihypertensive medications or SBP ≥140 mmHg and/or DBP ≥90 mmHg at two separate evaluations) before age 60, their unmedicated adult offspring, and age-matched controls. Later the study population was expanded to include other siblings of the original sibling pair as well as any offspring for a total sample size of N=5000. The Black participants underwent whole genome sequencing (WGS), through the National Heart Lung and Blood Institute (NHLBI) WGS program. In order to harmonize our imputation efforts with the array-based panels of the REGARDS, GenHAT and Warfarin (see below) studies, we compiled a set of non-monomorphic and non-multi-allelic SNPs with MAF >1% that were genotyped for GWAS as part of those studies. This yielded a total of 2,204,415 SNPs that were used as fence post markers for imputation. In order to maintain consistency HyperGEN samples were then imputed into the same version of TOPMed release2 reference panel that was adopted for imputing REGARDS, GenHAT and Warfarin. Imputed variants were inspected for their imputation quality scores (R^2^) and it was noted that more than 99% of the variants with MAF>1% had an imputation quality of 0.6 or higher. Given the high-quality of imputation for variants of MAF>1% and to retain maximum overlap of variants with the SNPs with PRS weights, genotypes with genotypic probability of 0.9 or higher were retained. Participants were excluded if they were younger than 40 years at enrollment, leaving 1,898 self-identified Black participants for analysis. *APOL1* genotypes were obtained directly from the WGS data.

#### Warfarin Pharmacogenomics Cohort (WPC)

WPC is a prospective cohort of first-time warfarin users aged 19 years or older starting warfarin for anticoagulation^65,66^. Warfarin therapy requiring a target international normalized ratio (INR) range of 2-3 was initiated in patients with venous thromboembolism, stroke/transient ischemic attacks, atrial fibrillation, myocardial infarction, and/or peripheral arterial disease. Patients requiring a higher intensity (INR 2.5 to 3.5) or lower intensity (INR 1.5 to 2.5) of anticoagulation were excluded. A detailed history was obtained that included baseline demographics, as well as medication history and compliance. Changes in INR, medications and laboratory parameters were documented at each clinical visit as reported previously. The WPC includes genotyped data using the Illumina MEGA-EX array, as well as an Illumina IM duo array for 599 and 297 Black participants, respectively. Imputation was performed using the NHLBI TOPMed r2 reference panel (Freeze 8). Imputed variants were inspected for their imputation quality scores (R^2^) and it was noted that more than 99% of the variants with MAF>1% had an imputation quality of 0.6 or higher. Given the high-quality of imputation for variants of MAF>1%, in order to retain maximum overlap of variants with the SNPs with PRS weights, genotypes with genotypic probability of 0.9 or higher were retained. This strategy retained highest-quality genotype calls for the SNPs that were employed for the PRS derivation. Ten principal components (PCs) were calculated using a program EIGENSOFT (version 6.1.4) selecting directly genotyped SNPs (MAF ≥ 5%, missing data < 5%, and HWE p-value >1E-04) based on pairwise linkage disequilibrium (LD, using r2 < 0.05). This resulted in 44,137 tagged SNPs in African American samples. *APOL1* information was obtained from genotypic array data; rs143830837 (bp hg38 36265995) was used as a proxy for rs71785313 (bp hg38 36265996) since these SNPs represent the same G2 variant and were recently merged in the NCBI dbSNP database (https://www.ncbi.nlm.nih.gov/snp/?term=rs143830837). For this analysis, only participants aged 40 years or older were included, leaving a total of 448 self-identified Black participants.

#### The Genetics of Hypertension Associated Treatments (GenHAT) Study

GenHAT is an ancillary study to the Antihypertensive and Lipid Lowering Treatment to Prevent Heart Attack Trial (ALLHAT)^67^. ALLHAT was a randomized, double blind, multicenter clinical trial with over 42,000 high-risk individuals with hypertension, aged 55 years or older, and had at least one additional risk factor for CVD. ALLHAT is the largest antihypertensive treatment trial to date and was ethnically diverse, enrolling over 15,000 Black participants^68^. Participants were randomized into four groups defined by the class of assigned antihypertensive medication including chlorthalidone, lisinopril, amlodipine, and doxazosin at a ratio of 1.7:1:1:1, respectively. The original GenHAT study (N=39,114) evaluated the effect of the interaction between candidate hypertensive genetic variants and different antihypertensive treatments on the risk of fatal and non-fatal CVD outcomes^67^. In an ancillary study to the original GenHAT study, participants self-identified as Black were genotyped for GWAS to better understand genes involved in response to chlorthalidone and lisinopril. Genotyping using Illumina MEGA-EX arrays was performed on 7,546 Black adults who met these criteria. Upon genotyping completion samples that failed or had low call rate (<95%) were excluded. Participants were also excluded with sex mismatches, or if they were an outlier on principal component analysis (outside of 6 standard deviations), which resulted in 6,919 Black GenHAT participants with genotypes available for analysis. Imputation was carried out using the NHLBI TOPMed release 2 (r2) reference panel (Freeze 8). Imputed variants were inspected for their imputation quality scores (R^2^) and more than 99% of the variants with MAF>1% had an imputation quality of 0.6 or higher. Given the high-quality of imputation for variants of MAF>1%, in order to retain maximum overlap of variants with the SNPs with PRS weights, genotypes with genotypic probability of 0.9 or higher were retained. *APOL1* information was extracted from the genotypic array data. Similar to WPC, rs143830837 variant was used as a proxy for the *APOL1* G2 allele.

### Ancestry definitions

In UKBB and eMERGE-III datasets, the ancestry sub-cohorts were defined based on clustering on principal component analysis of genetic markers. We grouped all individuals into major continental ancestry clusters: European, African, Hispanic/Latinx, South Asian, and East Asian. This was done by projecting each sample onto the reference principal components calculated from the 1000G reference panel^69^. Briefly, we merged our UKBB and eMERGE samples with 1000G samples and kept only SNPs in common between the two datasets. We used common variants between UKBB and eMERGE with 1000G following command in PLINK --indep-pairwise 500 50 0.05. The numbers of pruned variants for UKBB and eMERGE were 35,091 and 43,080 respectively. We then calculated principal components (PCs) for the 1000G samples using FlashPCA and projected each of our samples onto those PCs. Ancestry assignments were then performed by co-clustering of the 1000G reference populations and ancestry memberships were verified by visual inspection of PCA plots. Ancestry in BioMe, REGARDS, HyperGEN, WPC, and GenHAT was determined by self-reported race/ethnicity, and PCA was subsequently performed for verification and to exclude outliers.

### CKD phenotyping and case-control definitions

For CKD phenotyping based on EHR data, we used a computable CKD phenotype recently developed and extensively validated by the eMERGE-III network^4^. In the population-based UKBB datasets, we defined cases as those having CKD stage 3 or above (based on eGFR estimated with CKD-EPI equation^70^, and including patients on chronic dialysis or after a kidney transplant) and compared them to population controls. In all other case-control testing datasets, we defined cases using the same definition (CKD stage 3 or above based on CKD-EPI eGFR^70^, or patients on chronic dialysis or after a kidney transplant), while controls were defined as those without known CKD and eGFR > 90 mL/min/1.73m^2^ based on the latest serum Cr available. While reducing the overall sample size of our control datasets, the exclusion of individuals with CKD stage 2 (eGFR 60-90 mL/min/1.73m^2^) aimed to minimize any potential misclassification of the case-control status. Only individuals 40 years of age or older were included across all datasets for consistency with the UKBB ascertainment strategy. Additional covariates used in the predictive models included age, sex, diabetes (type I or II), and principal components of ancestry. The diagnosis of diabetes type I or II was added as an important covariate, given that it represents an established large-effect risk factor for CKD and kidney failure. The diagnosis of hypertension was not added as a covariate to avoid over-adjustment, since case-effect relationship of hypertension to CKD is usually not clear in electronic health records, and CKD itself represents the most common cause of secondary hypertension.

### Polygenic score design and optimization

We used 70% Europeans of UKBB (28,047 cases and 251,772 controls) to optimize the GWAS-based polygenic component of the GPS by selecting the best model between two commonly used methods and a range of input parameters (“ Optimization Dataset” , **Table 1, Supplementary Table 2**). We used the summary statistics for 8.2 million common variants from the CKDGen consortium GWAS for eGFR^17^ in combination with the diverse LD reference panel from phase 3 1000G project (all populations, N=2,504)^59^. We first computed 7 candidate GPSes using the LDPred computational algorithm^71^ across the following range of rho (fraction of casual variants): 1.00E+00, 1.00E-01, 1.00E-02, 1.00E-03, 3.00E-01, 3.00E-02 and 3.00E-03. We also generated 12 pruning and thresholding (P+T) scores with r^2^=0.2 and P-value thresholds of 1.0, 1.00E-02, 1.00E-03, 1.00E-04, 1.00E-05, 1.00E-06, 1.00E-07, 1.00E-08, 3.00E-02, 3.00E-03, 3.00E-04 and 3.00E-05. Based on the above parameters, each GPS was expressed as a weighted sum of alleles with weights based on the GWAS for the eGFR study:

**Table 1:** Summary of study cohorts used for GPS optimization and testing.

| Study | Sub-Cohort | CKD Cases<br>N=59,168 | Controls<br>N=398,363 | Female<br>(%) | Diabetes<br>(%) | Mean Age<br>(Years) |
| --- | --- | --- | --- | --- | --- | --- |
| UKBB | Optimization 1: European Ancestry (70%) | 28,047 | 251,772 | 46 | 5 | 56.65 |
|  | Optimization 2: African Ancestry | 967 | 6,191 | 42 | 12 | 51.77 |
|  | Testing 1: European Ancestry (30%) | 11,922 | 108,002 | 46 | 5 | 56.64 |
|  | Testing 2: East Asian Ancestry | 74 | 1,740 | 32 | 5 | 52.37 |
|  | Testing 3: South Asian Ancestry | 797 | 7,698 | 54 | 18 | 53.32 |
| eMERGE | Testing 4: European Ancestry | 10,572 | 8,030 | 48 | 35 | 71.23 |
|  | Testing 5: African Ancestry | 1,143 | 1,600 | 30 | 40 | 66.76 |
|  | Testing 6: Hispanic Ancestry | 488 | 639 | 37 | 36 | 67.75 |
|  | Testing 7: East Asian Ancestry | 98 | 105 | 39 | 26 | 72.27 |
| UAB | Testing 8: Warfarin: African Ancestry | 308 | 140 | 58 | 50 | 61.48 |
|  | Testing 9: REGARDS: African Ancestry | 1,055 | 4,314 | 63 | 31 | 62.30 |
|  | Testing 10: GenHAT: African Ancestry | 924 | 2,454 | 58 | 45 | 65.74 |
|  | Testing 11: HyperGEN: African Ancestry | 109 | 619 | 62 | 31 | 52.23 |
| BioMe | Testing 12: European Ancestry | 870 | 1,851 | 38 | 14 | 61.87 |
|  | Testing 13: African Ancestry | 729 | 1,149 | 56 | 32 | 61.65 |
|  | Testing 14: Hispanic Ancestry | 1,004 | 1,706 | 38 | 33 | 62.04 |
|  | Testing 15: East Asian Ancestry | 61 | 353 | 41 | 15 | 55.75 |

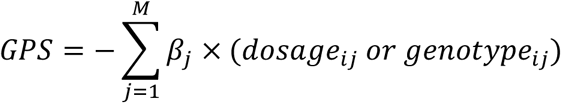

where M is number of variants in the model and *β*_*j*_ is the weight based on GWAS summary statistics and the negative sign reflects an inverse relationship between eGFR and CKD.

Each of the 19 scores derived above was subsequently assessed for discrimination of CKD cases from population controls in the first UKBB optimization dataset after adjustment for age, sex, diabetes status and four principal components of ancestry. The score with the best performance was defined by the maximal area under the receiver operator curve and the largest fraction of variance explained (**Supplementary Table 2**). The best performing score was normal-standardized (by subtracting control mean and dividing by control standard deviation) and advanced for testing in the second UKBB optimization cohort of African ancestry.

### Modeling the effects of *APOL1* risk genotypes

To optimize trans-ethnic performance, our final score was further optimized using the second, smaller UKBB optimization dataset of African ancestry (967 cases and 6,191 controls). We aimed to assess if adding *APOL1* risk genotype (under a recessive model) enhanced CKD risk prediction. For this purpose, we first removed any variants in the *APOL1* region from the GWAS-based GPS equation. Next, we tested the GPS and *APOL1* risk genotype jointly for association with CKD in this dataset. The GPS (without *APOL1* region) and recessive *APOL1* risk genotypes both represented independently significant predictors of CKD before and after adjustment for age, sex, diabetes (Type I or II), and 4 principal components of ancestry. The risk effects of *APOL1* and GPS were additive, with one SD unit of the standard-normalized GPS conveying the risk that was approximately equivalent to *APOL1* risk genotype (**Supplemental Table 3**). We also tested for effect modification of *APOL1* risk genotype by the polygenic component in CKD prediction, but we detected no significant interactive effects (P interaction = 0.29). To best account for an independent additive effect of recessive *APOL1* risk genotypes, we therefore updated the CKD GPS for each subject using the following equation:

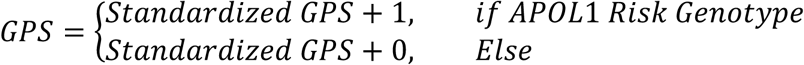

### Predictive performance in independent testing datasets

The predictive performance of the final risk score formulation was assessed in 15 ancestrally diverse testing datasets, including 3 cohorts of European ancestry (by genetic or self-reported European ancestry, 23,364 cases and 117,883 controls in total), 6 cohorts of African ancestry (4,268 cases and 10,276 controls), 4 cohorts of Asian (East and South-West) ancestry (1,030 cases and 9,896 controls), and 2 Hispanic/Latinx ancestry cohorts (1,492 cases and 2,984 controls). We calculated a full set of standardized risk score performance metrics following eMERGE-IV and ClinGen guidelines^48^. Logistic regression models were used for predicting case-control status with adjustment for age, sex, diabetes (Type I or II), center and genotype/imputation batch (if relevant), and four principal components of ancestry using glm function in R.

We used pROC R package to calculate the receiver operating characteristics area under curve (AUC), using logistic regression model with CKD case status as an outcome and the GPS, age, sex, center, batch, and four significant PCs as predictors. We calculated variance explained using the Nagelkerke’s pseudo-R^2^, including for the full model (GPS plus covariates), for the covariates-only model, and for the GPS component alone expressed as the R^2^ difference between the full and the covariates-only model. We also expressed the effect of standardized risk score as odds ratios (with 95% confidence intervals) per standard deviation unit of the control standard normalized risk score distribution in each of the validation cohorts. We examined the risk score discrimination at tail cut-offs corresponding to the top 20%, 10%, 5%, 2%, 1% of the GPS distribution by deriving odds ratios of disease for each tail of the distribution compared to all other individuals in each cohort. We also calculated sensitivities and specificities for each cut-off point in each testing cohort.

The performance metrics were then meta-analyzed across ancestry-defined testing cohorts using an inverse variance weighed fixed-effects method to derive pooled performance metrics for each ancestral grouping^72^. Finally, we calculated ancestry-specific prevalence-adjusted positive and negative predictive values for each GPS cut-off based on pooled estimates of sensitivity and specificity and known CKD prevalence in US population (https://www.cdc.gov/kidneydisease/publications-resources/ckd-national-facts.html). Statistical analyses were conducted using R version 3.6.3 (2019-02-29) software.

### Comparing GPS distributions in the 1000G reference populations

To assess differences in the distributions of GPS by ancestry, we computed risk scores for the multiethnic reference of all 1000G phase 3 participants using our final optimized CKD GPS equation:

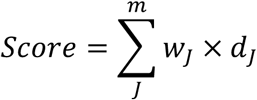

where *w*_*J*_ are optimized weights from CKD GWAS summary statistics for each marker included in the score, *d*_*J*_ is dosage or genotypes (0, 1 and 2) for 1000G samples and m is the total number of variants from GWAS summary statistics included in our final GPS (m=471,316). The distributions were examined visually in the form of histograms, and distributional differences by ancestry were tested using ANOVA.

### Post-hoc ancestry adjustment

In order to express GPS effects on the same scale across ancestrally diverse individuals and select a single cut-off for clinical implementation of the GPS, we adjusted for differences in the first two moments of the GPS distributions by ancestry. Using multiethnic eMERGE cohorts, we tested two different regression-based genetic ancestry adjustment strategies that utilize 1000G (all populations) reference: **method 1** which adjusts for differences in mean and **method 2** which adjusts for both differences in mean and variance.

For **method 1**, we first regressed the GPS of 1000G participants against the first five PCs as proposed previously^73^:

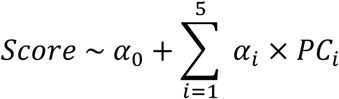

Fitting the model to 1000G reference panel allows us to find *α*’s and generate residuals. Next, we used the predicated *α*’s to calculate the adjusted score for any individual projected onto the same PCA space:

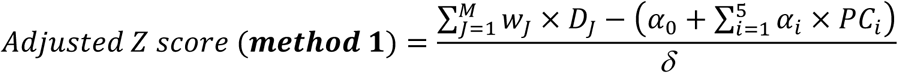

where 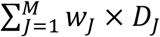 is the raw GPS, 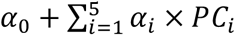 is the predicted (ancestry-adjusted) mean, and *δ* is the residual standard deviation from the 1000G model (all populations).

To adjust for ancestral differences in both mean and variance (**method 2**), we used the same method as above, but we also modeled residual variance (*δ*^2^) as a function of PCs of ancestry:

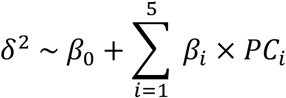

Next, we used the predicated *α* ‘s and *β*’s to calculate the adjusted Zp score:

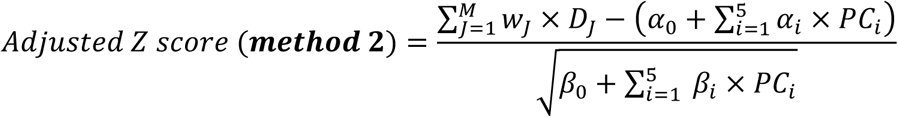

Where 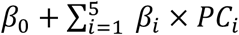 sis the predicted (ancestry-adjusted) residual variance.

The distributional transformations achieved by these methods were examined visually. We then compared the effects of these adjustments for the top percentile cut-offs in eMERGE-III cohorts. We also assessed the overall GPS calibration across all eMERGE cohorts after final ancestry adjustment.

## RESULTS

### GPS Optimization

The flowchart summary of the GPS derivation and testing strategy is provided in **Figure 1**. In the optimization step, a total of 19 candidate risk scores were generated based on multiethnic LD reference panel and summary statistics from GWAS for eGFR^17^. We then used a large optimization dataset comprised of 70% of European UK Biobank participants to select the best performing model (**Table 1, Supplementary Table 2**). The best model was based on the P+T method and involved 471,316 markers selected based on r2 = 0.2 and P < 0.03. The score was standardized to zero-mean and unit-variance based on ancestry-matched population controls. In the optimization dataset, the polygenic component of the risk score explained 3% of variance (R^2^), with one standard deviation of the score increasing CKD risk by 60% (OR=1.60, 95%CI 1.59-1.61, P <1.00E-300) after controlling for age, sex, diabetes, genotyping batch, and genetic ancestry (**Supplementary Table 2**).

**Figure 1:**
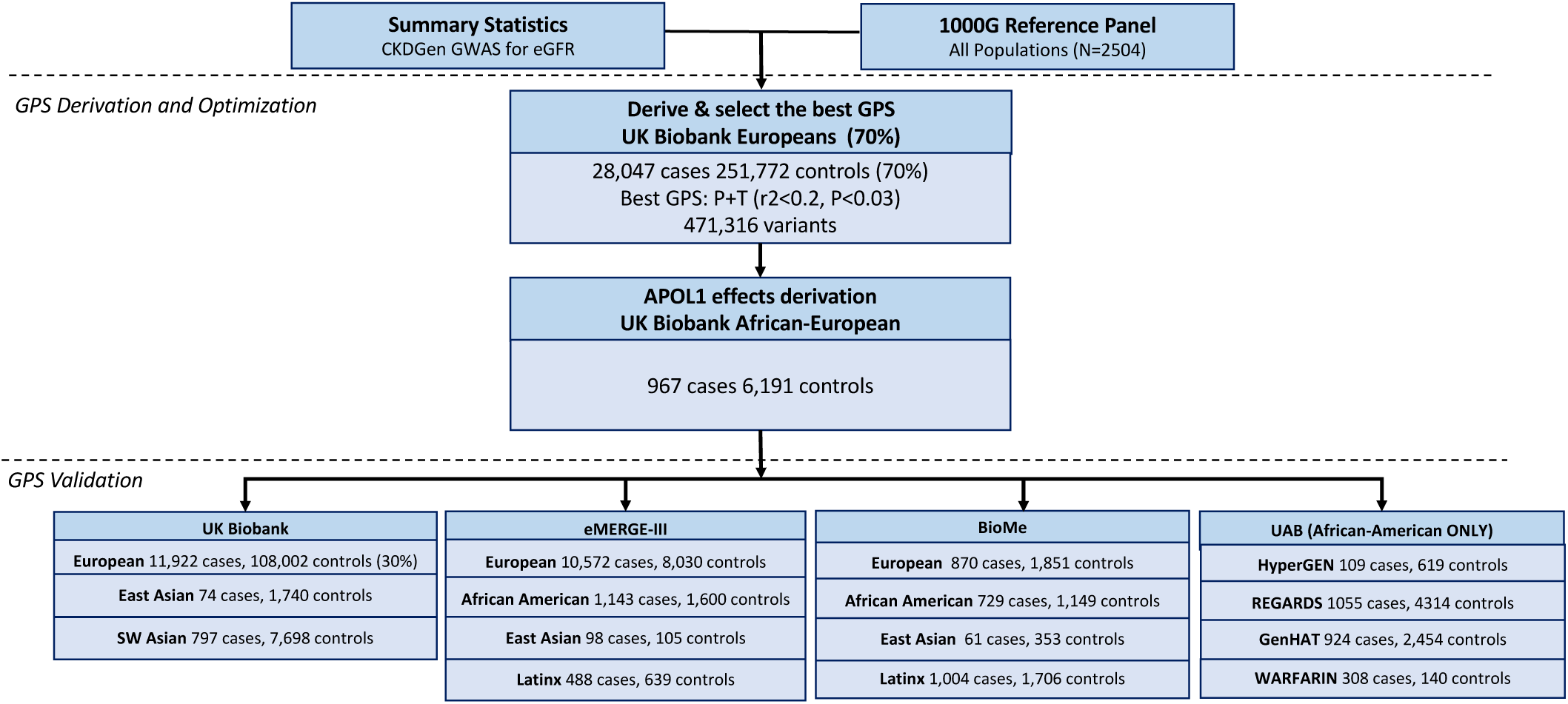
Overview of the study design. The CKD GPS was designed based on CKDGen GWAS summary statistics for eGFR and a cosmopolitan LD reference panel of 1000 Genomes (all populations); optimization was performed in two stages using UKBB participants of European (optimization 1) and African (optimization 2) ancestries; GPS performance validation was conducted in 15 additional independent testing cohorts of diverse ancestries.

The second optimization step involved testing for independent contributions of *APOL1* risk genotypes and included 7,158 UKBB participants of genetically defined African ancestry (967 cases and 6,191 controls). In the model adjusted for age, sex, diabetes (Type I and II), batch, and PCs of ancestry, we observed statistically significant independent effects of the polygenic component (OR per SD =1.16, 95%CI: 1.09-1.25, P=1.00E-04) and the recessive *APOL1* risk genotype (OR=1.19, 95%CI: 1.01-1.38, P=4.00E-02), but no significant multiplicative interactions between the two predictors (P interaction=0.29) (**Supplementary Table 3**). Given these findings, we subsequently modeled *APOL1* risk as additive to the polygenic component, with the recessive risk genotype effects approximately equivalent to one standard deviation of the standard normalized polygenic score (a weight of one standard deviation unit was selected because the β per standard deviation of the polygenic score and the β for APOL1 risk genotype were comparable in magnitude).

### Population differences in GPS distributions

We next examined the distributions of the polygenic risk component (without *APOL1*), as well as the final combined GPS (with *APOL1*) in the reference populations of 1000 Genomes. We detected significant differences in the mean polygenic risk across reference populations (**Figure 2**, ANOVA P=3.40E-154), with a notable shift towards higher average risk in the African population compared to all other populations (P=4.92E-163). This shift became even more pronounced after the inclusion of *APOL1* risk genotype information in the combined GPS (P=1.58E-168). These results suggest that the polygenic risk for CKD is considerably higher in African compared to non-African populations independent of *APOL1*.

**Figure 2:**
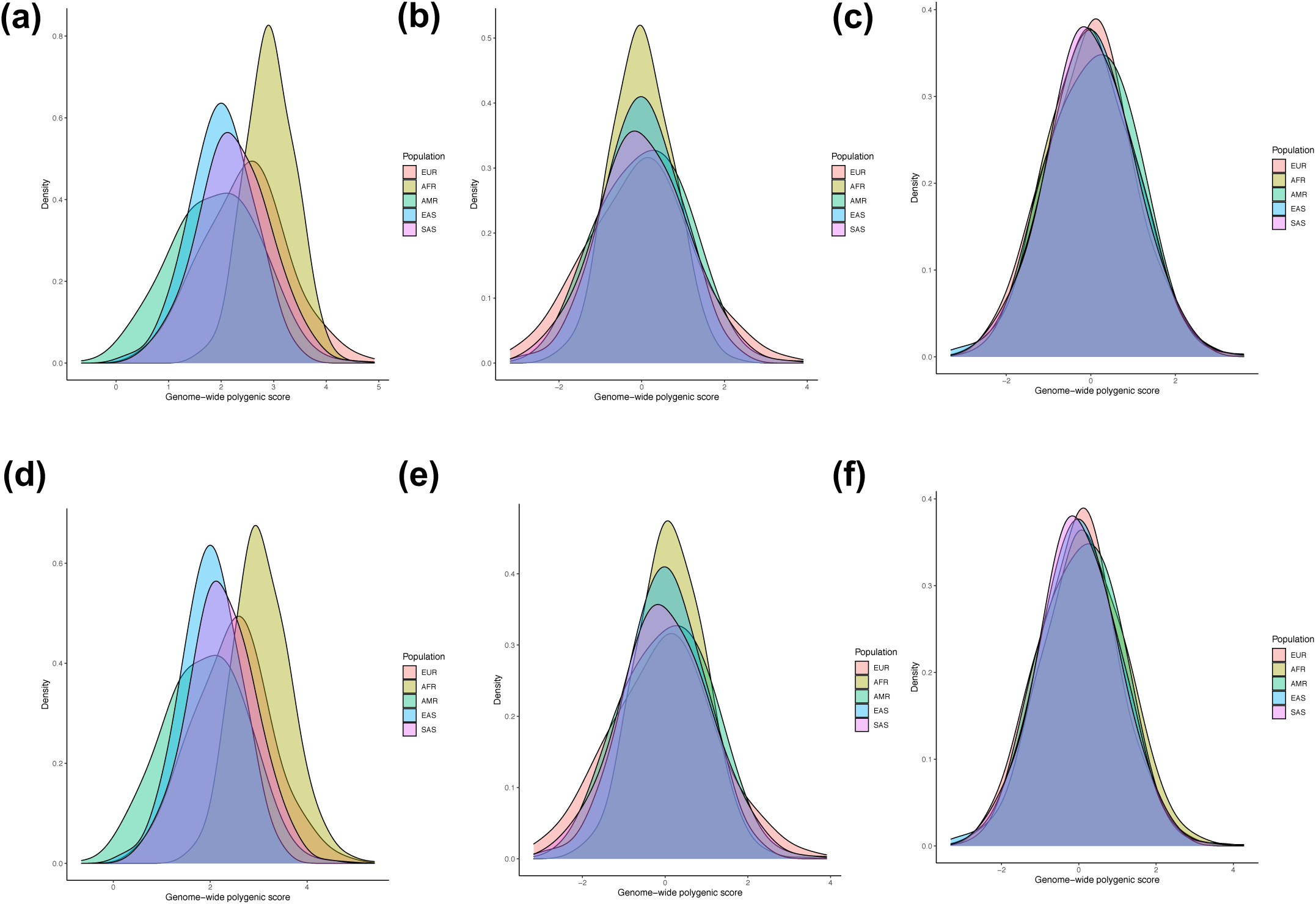
Risk score distributions in five 1000 Genomes populations: **(a)** raw polygenic score without *APOL1*; **(b)** ancestry-adjusted polygenic score without *APOL1* (method 1: mean only); **(c)** ancestry-adjusted polygenic score without *APOL1* (method 2: mean and variance); **(d)** raw combined GPS with *APOL1*; **(e)** ancestry-adjusted combined GPS with *APOL1* (method 1) and **(f)** ancestry-adjusted combined GPS with *APOL1* (method 2). AFR: African, AMR: Native American, EAS: East Asian, EUR: European, and SAS: South Asian.

Given that the weights of the polygenic score equation are fixed and derived based on the GWAS performed predominantly in European cohorts, we hypothesized that these distributional differences are likely driven by a higher frequency of CKD risk alleles in African genomes. Therefore, we examined the overall frequency spectrum of CKD risk alleles included in the GPS between European and African reference populations (**Supplemental Figure S2**). As expected for variants selected based on European GWAS, we observed a greater number of risk alleles at the extremes of the frequency spectrum (RAF < 0.01 or > 0.099) in the African compared to European populations of 1000G. This observation is likely due to the routine use of MAF filter of 0.01 in the individual European GWAS discovery cohorts contributed to the GWAS meta-analysis for eGFR. Across all variants included in the score, the mean difference in risk allele frequencies (RAF) between African and European populations was positive (i.e. greater than the expected mean=0), and this rightward shift in the distribution of RAF difference is indicative of higher average frequency of risk alleles in African genomes. We further observed that the risk alleles with largest weights (effect sizes in GWAS) had a significantly higher frequency in African genomes compared to those with low effect sizes (P=0.02), or intermediate effect sizes (P=0.018) (**Supplemental Figure S2d)**. Thus, it appears that the observed GPS distributional shifts between European and African populations are driven predominantly by frequency differences of large effect risk alleles.

### GPS Testing in cohorts of European ancestry

We next tested the final GPS in three European cohorts, including the remaining 30% of the UKBB (11,922 cases and 108,002 controls) and two large US-based European ancestry cohorts, eMERGE-III (10,572 cases and 8,030 controls) and Bio*Me* (870 cases and 1,851 controls). In the combined meta-analysis across the three testing cohorts, the GPS exhibited highly reproducible performance, with pooled OR per SD = 1.49, 95%CI: 1.47-1.50, P < 1.00E-300 and ROC AUC=0.75, 95%CI: 0.75-0.76 (**Supplemental Table 4**). While the UKBB testing cohort had nearly identical performance metrics to the UKBB optimization cohort (OR per SD = 1.60, 95%CI: 1.58-1.62, P < 1.00E-300) the effect sizes were slightly attenuated in the US-based cohorts (eMERGE-III: OR per SD=1.38, 95%CI:1.35-1.40, P=1.58E-83 and Bio*Me*: OR per SD=1.58, 95%CI:1.46-1.70, P=2.50E-14. We also note that the frequency of *APOL1* risk genotype was extremely low, thus its effect was negligible in European cohorts.

### GPS testing in cohorts of African ancestry

The GPS was tested in six independent African ancestry cohorts, including eMERGE-III (1,143 cases and 1,600 controls), Bio*Me* (729 cases and 1,149 controls), HyperGEN (109 cases and 619 controls), REGARDS (1,055 cases and 4,314 controls), GenHat (924 cases and 2,454 controls) and Warfarin (308 cases and 140 controls). In the combined meta-analysis, the GPS had pooled OR per SD = 1.32, 95%CI: 1.26-1.38, P =1.77E-33 and ROC AUC=0.78, 95%CI: 0.77-0.79 (**Table 2 and Supplemental Table 5**). We note that the inclusion of *APOL1* risk genotype in the GPS considerably enhanced CKD risk prediction across all cohorts of African ancestry, substantially improving tail cut-off discrimination i.e., the risk for the top 2% was approximately 1.8-fold higher in the model without *APOL1* and 2.7-fold higher in the model with APOL1, when compared to the remaining 98% of individuals (**Table 3)**.

**Table 2:**
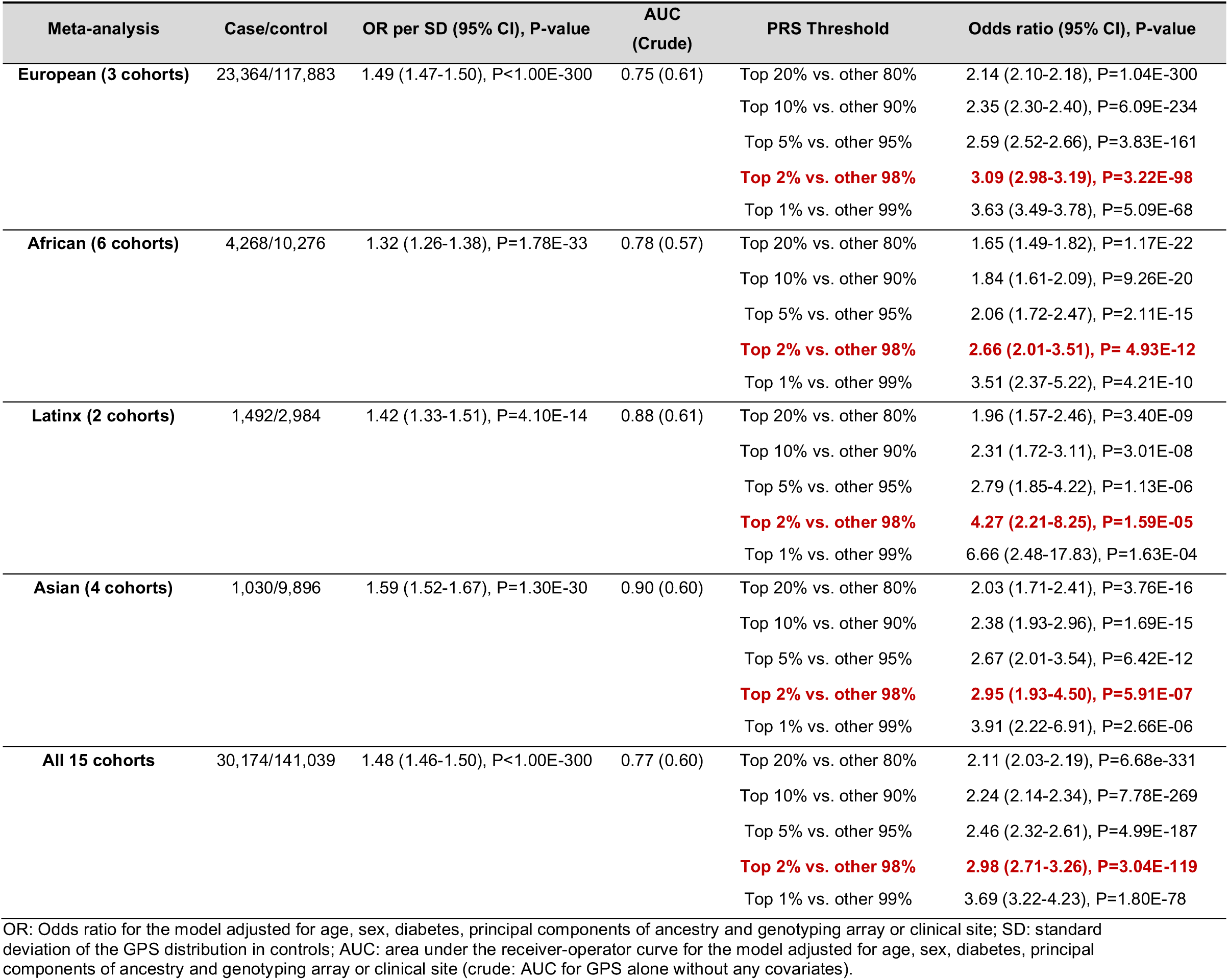
The performance metrics of the GPS in the testing cohorts meta-analyzed by continental ancestry. For performance testing in individual cohorts, please refer to Supplemental Tables 4-7.

| Meta-analysis | Case/control | OR per SD (95% CI), P-value | AUC (Crude) | PRS Threshold | Odds ratio (95% CI), P-value |
| --- | --- | --- | --- | --- | --- |
| **European (3 cohorts)** | 23,364/117,883 | 1.49 (1.47-1.50), P<1.00E-300 | 0.75 (0.61) | Top 20% vs. other 80% | 2.14 (2.10-2.18), P=1.04E-300 |
|  |  |  |  | Top 10% vs. other 90% | 2.35 (2.30-2.40), P=6.09E-234 |
|  |  |  |  | Top 5% vs. other 95% | 2.59 (2.52-2.66), P=3.83E-161 |
|  |  |  |  | **Top 2% vs. other 98%** | **3.09 (2.98-3.19), P=3.22E-98** |
|  |  |  |  | Top 1% vs. other 99% | 3.63 (3.49-3.78), P=5.09E-68 |
| **African (6 cohorts)** | 4,268/10,276 | 1.32 (1.26-1.38), P=1.78E-33 | 0.78 (0.57) | Top 20% vs. other 80% | 1.65 (1.49-1.82), P=1.17E-22 |
|  |  |  |  | Top 10% vs. other 90% | 1.84 (1.61-2.09), P=9.26E-20 |
|  |  |  |  | Top 5% vs. other 95% | 2.06 (1.72-2.47), P=2.11E-15 |
|  |  |  |  | **Top 2% vs. other 98%** | **2.66 (2.01-3.51), P= 4.93E-12** |
|  |  |  |  | Top 1% vs. other 99% | 3.51 (2.37-5.22), P=4.21E-10 |
| **Latinx (2 cohorts)** | 1,492/2,984 | 1.42 (1.33-1.51), P=4.10E-14 | 0.88 (0.61) | Top 20% vs. other 80% | 1.96 (1.57-2.46), P=3.40E-09 |
|  |  |  |  | Top 10% vs. other 90% | 2.31 (1.72-3.11), P=3.01E-08 |
|  |  |  |  | Top 5% vs. other 95% | 2.79 (1.85-4.22), P=1.13E-06 |
|  |  |  |  | **Top 2% vs. other 98%** | **4.27 (2.21-8.25), P=1.59E-05** |
|  |  |  |  | Top 1% vs. other 99% | 6.66 (2.48-17.83), P=1.63E-04 |
| **Asian (4 cohorts)** | 1,030/9,896 | 1.59 (1.52-1.67), P=1.30E-30 | 0.90 (0.60) | Top 20% vs. other 80% | 2.03 (1.71-2.41), P=3.76E-16 |
|  |  |  |  | Top 10% vs. other 90% | 2.38 (1.93-2.96), P=1.69E-15 |
|  |  |  |  | Top 5% vs. other 95% | 2.67 (2.01-3.54), P=6.42E-12 |
|  |  |  |  | **Top 2% vs. other 98%** | **2.95 (1.93-4.50), P=5.91E-07** |
|  |  |  |  | Top 1% vs. other 99% | 3.91 (2.22-6.91), P=2.66E-06 |
| **All 15 cohorts** | 30,174/141,039 | 1.48 (1.46-1.50), P<1.00E-300 | 0.77 (0.60) | Top 20% vs. other 80% | 2.11 (2.03-2.19), P=6.68e-331 |
|  |  |  |  | Top 10% vs. other 90% | 2.24 (2.14-2.34), P=7.78E-269 |
|  |  |  |  | Top 5% vs. other 95% | 2.46 (2.32-2.61), P=4.99E-187 |
|  |  |  |  | **Top 2% vs. other 98%** | **2.98 (2.71-3.26), P=3.04E-119** |
|  |  |  |  | Top 1% vs. other 99% | 3.69 (3.22-4.23), P=1.80E-78 |

OR: Odds ratio for the model adjusted for age, sex, diabetes, principal components of ancestry and genotyping array or clinical site; SD: standard deviation of the GPS distribution in controls; AUC: area under the receiver-operator curve for the model adjusted for age, sex, diabetes, principal components of ancestry and genotyping array or clinical site (crude: AUC for GPS alone without any covariates).

**Table 3:** Added value *APOL1* risk genotype to polygenic risk components in predicting CKD using extreme tail (98^th^ percentile) of the risk score distribution in African-American (4,268 cases and 10,276 controls) and Hispanic (1,492 cases and 2,984 controls) cohorts. All effect estimates are adjusted for age, sex, diabetes, and principal components of ancestry.

| Cohorts | *APOL1* Risk Genotype<br>OR (95%CI), P-value | Top 2% PRS without *APOL1*<br>OR (95%CI), P-value | Top 2% PRS with *APOL1*<br>OR (95%CI), P-value |
| --- | --- | --- | --- |
| **African Ancestry:** |  |  |  |
| eMERGE | 1.64 (1.42-1.86), P=2.0E-05 | 2.10 (1.46-2.74), P=2.0E-02 | 2.60 (1.38-4.90), P=3.10E-03 |
| BioMe | 1.38 (1.28-1.48), P=3.3E-10 | 2.70 (1.93-3.47), P=1.0E-02 | 5.75 (4.96-6.54), P=1.00E-05 |
| UAB HyperGen | 1.71 (0.93-3.12), P=8.2E-02 | 2.22 (0.59-8.44), P=2.4E-01 | 1.64 (0.43-6.20), P=4.65E-01 |
| UAB REGARDS | 1.35 (1.08-1.77), P=6.9E-03 | 1.26 (0.76-2.07), P=3.6E-01 | 1.56 (0.97-2.59), P=6.52E-02 |
| UAB GenHAT | 1.43 (1.12-1.81), P=3.2E-03 | 2.80 (1.64-4.77), P=1.5E-04 | 4.38 (2.56-7.50), P=6.80E-08 |
| UAB Warfarin | 1.93 (1.07-3.49), P=2.9E-02 | -- | 1.59 (0.28-8.73), P=5.96E-01 |
| **Meta-analysis** | **1.46 (1.38-1.54), P=2.7E-19** | **1.76 (1.41-2.20), P=5.9-07** | **2.66 (2.01-3.51), P=4.93E-12** |
| **Hispanic Ancestry:** |  |  |  |
| eMERGE | 15.8 (3.11-80.9), P=9.1E-04 | 1.58 (0.55-2.60), P=3.8E-01 | 3.82 (1.15-12.62), P=2.83E-02 |
| BioMe | 1.17 (1.10-1.24), P=2.4E-06 | 2.72 (1.97-3.47), P=1.0E-02 | 4.48 (3.69-5.27), P=2.10E-04 |
| **Meta-analysis** | **1.18 (1.09-1.26), P=4.5E-05** | **2.72 (2.19-3.25), P=2.0E-04** | **4.27 (2.21-8.25), P=1.59E-05** |

### GPS testing in cohorts of Hispanic/Latinx ancestry

The GPS was also tested in two Hispanic/Latinx cohorts from eMERGE-III (488 cases and 639 controls) and Bio*Me* (1,004 cases and 2,345 controls). The combined meta-analysis of these cohorts resulted in pooled OR per SD = 1.42, 95%CI: 1.33-1.51, P=4.10E-14 and ROC AUC 0.88, 95%CI: 0.85-0.89 (**Supplementary Table 6)**. As expected, due to high rates of African admixture in the Latinx population, the inclusion of *APOL1* risk genotypes in the GPS also improved predictive performance and tail discrimination of the GPS in these cohorts (**Table 3**).

### GPS testing in cohorts of Asian ancestry

Lastly, we evaluated GPS performance in four diverse Asian cohorts including UKBB South Asian (797 cases and 7,698 controls), UKBB East Asian (74 cases and 1,740 controls), eMERGE-III East Asian (98 cases and 105 controls) and Bio*Me* East Asian (61 cases and 353 controls) cohorts. The combined meta-analysis revealed results that were very similar to Europeans, with pooled OR per SD = 1.59, 95%CI: 1.52-1.67, P =1.30E-30 and ROC AUC=0.90, 95%CI: 0.87-0.92 (**Supplementary Table 7**). *APOL1* risk genotypes were absent in the Asian cohorts, thus the modelled risk was entirely attributable to the polygenic component.

### Tail discrimination performance by ancestry

For each individual testing cohort, we derived risk estimates comparing extreme tails of the risk score distribution to all other cohort members, and estimated sensitivity and specificity for a range of tail cutoffs (20%, 10%, 5%, 2% and 1%). These metrics were meta-analyzed by ancestry and are summarized in **Table 2 and Supplemental Tables 4-7**. Depending on ancestry, the top 2% tail of the risk score distribution was associated with 2-4 fold higher risk of CKD than the remaining 98% of individuals, including European (OR=3.09, 95%CI: 2.98-3.19, P=3.22E-98), African (OR=2.66, 95%CI: 2.01-3.51, P=4.93E-12), Latinx (OR=4.27, 95%CI: 2.21-8.25, P=1.59E-05) and Asian (OR=2.95, 95%CI: 1.93-4.50, P=5.91E-07) cohorts. We have therefore selected this cut-off as potentially clinically meaningful, since this degree of risk is approximately equivalent to the risk conferred by a positive family history of kidney disease. In **Table 4**, we summarize various metrics of diagnostic performance for this cut-off by ancestry, including sensitivity, specificity, as well as prevalence-adjusted positive and negative predictive values. For comparison, we also provide similar metrics for the cut-offs of top 5% of the risk score distribution in controls.

**Table 4:**
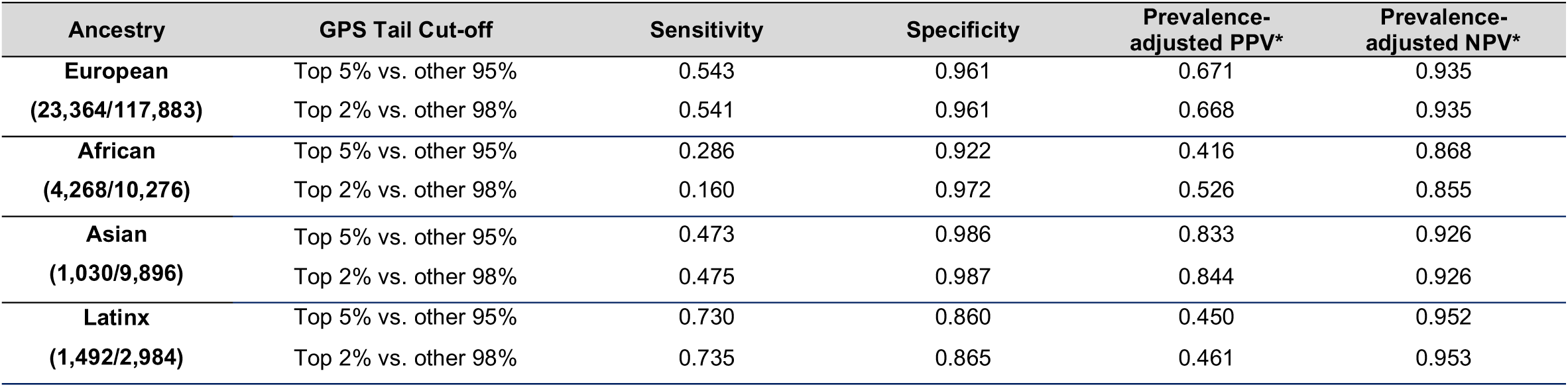
Pooled sensitivity, specificity, and prevalence-adjusted PPV and NPV for extreme tail cut-offs (95^th^ and 98^th^ percentile) of the GPS distribution in controls. The CKD prevalence was assumed to be 12.7% in European Americans; 16.3% in African Americans; 13.60% in Hispanic Americans, and 12.9% in Asian Americans as per the most recent CDC data (2021).

| Ancestry | GPS Tail Cut-off | Sensitivity | Specificity | Prevalence-adjusted PPV* | Prevalence-adjusted NPV* |
| --- | --- | --- | --- | --- | --- |
| **European**<br>(23,364/117,883) | Top 5% vs. other 95% | 0.543 | 0.961 | 0.671 | 0.935 |
|  | Top 2% vs. other 98% | 0.541 | 0.961 | 0.668 | 0.935 |
| **African**<br>(4,268/10,276) | Top 5% vs. other 95% | 0.286 | 0.922 | 0.416 | 0.868 |
|  | Top 2% vs. other 98% | 0.160 | 0.972 | 0.526 | 0.855 |
| **Asian**<br>(1,030/9,896) | Top 5% vs. other 95% | 0.473 | 0.986 | 0.833 | 0.926 |
|  | Top 2% vs. other 98% | 0.475 | 0.987 | 0.844 | 0.926 |
| **Latinx**<br>(1,492/2,984) | Top 5% vs. other 95% | 0.730 | 0.860 | 0.450 | 0.952 |
|  | Top 2% vs. other 98% | 0.735 | 0.865 | 0.461 | 0.953 |

### Ancestry adjustments and GPS calibration

We next compared the effect of two different ancestry adjustment methods on the GPS distributions in 1000G, eMERGE-III, and UKBB testing cohorts (**Figure 2 and Supplemental Figure S1**). Adjusting for mean only (**method 1**, see *methods*) eliminated major distributional shifts by ancestry, but did not fully resolve the observed tail differences. The ancestry adjustment **method 2** (adjusting for both mean and variance) results in comparable shapes of the GPS distributions by ancestry and facilitates the selection of a single trans-ancestry cut-off to define a “ high risk” group. Both methods result in comparably good risk score calibration when applied to the combined multiethnic eMERGE-III dataset (**Supplementary Figure S3)**. As a trade-off, however, the ancestry adjustment methods reduce tail discrimination at extreme cut-offs as summarized **Supplementary Table 8**. This trade-off appears most pronounced for **method 2** and for more admixed cohorts (African-American and Latinx).

### Comparison with existing studies

While our manuscript was in preparation, an alternative version of a polygenic score for renal function was published by Yu et al.^74^ This score was based on the combined GWAS meta-analysis for eGFR involving the CKDGen consortium study^17^ and 90% of the UK Biobank, resulting in larger sample size but also higher proportion of Europeans (82%) contributing to summary statistics. Notably, the GPS designed by Yu et al. did not include *APOL1* risk genotype and was not optimized for transethnic prediction of CKD. The key design and validation differences between our GPS and the one by Yu et al. are summarized in **Supplementary Table 9**.

Because the score by Yu et al. has not been tested in multi-ethic cohorts, we next tested its performance in our independent eMERGE-III cohorts (**Supplementary Table 10**). As expected given the design differences, the score by Yu et al. performed better in cohorts of European ancestry (R^2^ 3.5% vs. 2.2%), comparably well in Hispanic cohorts (R^2^ 1.2% vs. 1.1%), but was less predictive in cohorts of African (R^2^ 1.0% vs. 1.4%) or East Asian ancestry (R^2^ 1.0% vs. 2.6%). In the African ancestry testing cohort, individuals in the top 2% of the GPS distribution by Yu et al. had 92% increase in the risk of CKD (OR 1.92, 95%CI: 1.03-3.59, P=0.039), compared to 2.6-fold increased risk using the GPS developed in this study (OR 2.60, 95%CI: 1.38-4.90, P=0.0031). Lastly, we note that the African ancestry distribution of the GPS by Yu et al. was also shifted towards higher values in compared to other ancestries (**Supplementary Figure 4**), confirming that the observed distributional shift is independent of any specific method used to design the score.

## DISCUSSION

We developed a genome-wide polygenic score for CKD that captures the most recent data on the polygenic determinants of renal function, predicts CKD across four major continental ancestry groups, and is transferable across various genotyping platforms and imputation methods. We also developed a continuous ancestry adjustment to allow for trans-ancestry standardization of the risk score. Our score was designed and validated following the ClinGen and the PRS Catalogue guidelines and can be implemented as a clinical test for individual level risk prediction^48^. Importantly, our testing studies demonstrated that extreme tails of the risk score distribution (top 2%) convey over 2-3-fold increase in the disease risk across the four major continental ancestries. From the clinical perspective, this magnitude of risk could be considered as actionable given its equivalence to a positive family history of kidney disease^75^.

Although our risk score is based on a multiethnic GWAS for eGFR, we recognize that the allelic effects used to develop our GPS are heavily biased by the predominant Euro-Asian composition of the discovery GWAS, including approximately 75% European, 23% East Asian, 2% African, and only <1% Hispanic ancestry participants. Because there are currently no studies of similar size performed in African American and Hispanic participants that could be used to improve the accuracy of effect estimates in these populations, our model assumes fixed allelic effects across all ancestral groups.

Nevertheless, we used a diverse linkage disequilibrium reference panel in order to improve the trans-ethnic performance of the score, and we further enhanced the model by including African ancestry-specific recessive *APOL1* risk genotypes known to have large effects on the risk of kidney disease. We demonstrated that *APOL1* risk genotypes (coded under a recessive model) have an additive effect with the polygenic component, and significantly enhance case-control discrimination in cohorts of African and Hispanic ancestry. We additionally tested two post-hoc ancestry corrections that improve comparability of the scores across diverse populations.

Several important limitations of this work need to be discussed. First, we are most limited by the lack of large-scale GWAS for renal function in non-European populations, as well as small size of existing cohorts that could be used for performance optimization in non-Europeans^76^. As a result, the largest cohorts presently available for robust risk score optimization are also of predominantly European ancestry. The assumption of fixed allelic effects across different ancestral groups is likely inaccurate, because many disease-related lifestyle factors and environmental exposures correlated with ancestry could modify allelic effects. Although it is not possible to overcome this limitation in the present study, our GPS approach could be enhanced in the future by inclusion of larger non-European GWAS studies for eGFR or CKD.

Second, the performance comparisons between different ancestral groups could be biased by differences in genotyping platforms and ascertainment methods employed by various biobanks and studies used for testing. For example, the eMERGE-III and Bio*Me* cohorts are ascertained among patients receiving care at tertiary medical centers in the US and are enriched in diseased individuals attending subspecialty clinics. As a result, these cohorts have higher rates of CKD and comorbidities when compared to population-based studies, such as UKBB.

Third, the ancestry definitions varied in our testing cohorts. For eMERGE and UKBB, the ancestry was defined agnostically using genetic approaches. In contrast, our analysis of the Bio*Me*, REGARDS, GenHAT, HyperGEN, and WPC cohorts relied on self-report. Despite these cohort differences, the risk score performance was similar in most testing cohorts regardless of the specific ancestry definition employed to define testing cohort.

Fourth, by design, our score models polygenic effects from GWAS for renal function as approximated by estimated GFR from serum Cr (filtration biomarker) rather than CKD itself. We recognize multiple limitations to the use of estimated GFR as a phenotype in GWAS, including the fact that serum Cr level is influenced by the rate of Cr production and metabolism in addition to kidney clearance. Accordingly, we designed the score to predict moderately advanced CKD (stage 3 and above) rather than a mild degree of renal dysfunction, to capture a clinically meaningful disease state. Notably, our risk score does not incorporate available information on the polygenic determination of albuminuria^77^ or primary kidney diseases^78,79^. However, GWAS for these traits that are published to date remain several orders of magnitude smaller in sample size compared to the GWAS for eGFR and thus incorporation of such data must await more powerful studies.

Fifth, we observed significant differences in the mean and variance of the GPS distributions by ancestry. The observed shift in the mean GPS towards higher values in individuals of African ancestry is independent of *APOL1* and is driven by a higher average RAF in the African genomes. The inter-population RAF differences are greatest for the risk alleles with largest effects. This pattern may be consistent with polygenic adaptation, but the effects of uncorrected population stratification in the discovery GWAS may also potentially explain this phenomenon^80^.

We note that the observed differences in the GPS distributions by ancestry represent a significant challenge for the clinical implementation of polygenic risk scores. The key problem is that it is not possible to select a single GPS threshold for all ancestries that results in the similar magnitude of risk. Therefore, we have explored several approaches that could be used to overcome this issue. One approach involves classifying anyone undergoing GPS testing into one of the four continental ancestry groups based on self-report, then using ancestry-specific risk score cut-offs to interpret the results. However, because of a potential for racial bias, the use of race in clinical algorithms has been discouraged.^81^ One could also classify a tested individual based on inferred genetic ancestry from SNP data and subsequently apply ancestry-specific cut-offs. This approach still categorizes individuals into distinct groups and can be inaccurate, especially for those individuals with highly admixed genomes. We have therefore tested two different regression-based ancestry correction methods that model a continuous spectrum of genetic ancestry based on the diverse reference panel of 1000 Genomes. We demonstrate that the reference population-based correction for both mean and variance can best align distribution tails for the purpose of selecting a single trans-ancestry cut-off to define individuals with comparable risk of CKD. This however, results in some performance trade-offs, especially in admixed populations. Although imperfect, this ancestry adjustment may be helpful in improving risk score standardization for clinical use in diverse populations.

In summary, we derived, optimized, and tested a new GPS for CKD across major ancestries and suggested new methods for its trans-ethnic standardization. We demonstrated that the effects of the polygenic component and *APOL1* risk genotypes on the risk of CKD are additive. We also observed that the polygenic component of the score is significantly higher in individuals of African ancestry compared to other ancestral groups. Our study also demonstrated that individuals in the highest 2% of the risk score distribution have nearly 3-fold increase in the disease risk, the degree of relative risk that is equivalent to a positive family history of kidney disease. Our results suggest that at the extreme tail cutoff of the GPS may provide a clinically actionable genetic test for screening and early detection of CKD. Other potential applications of the GPS may include improved risk stratification of potential living kidney donors, or enhanced quality assessment of deceased donor kidneys in the setting of transplant. The clinical utility of the GPS in these clinical settings will require further testing in large prospective studies.

### Data sharing: GPS catalogue deposition

The final formulation of the GPS for CKD along with the standardized metrics of performance were deposited in the GPS catalogue, accession number pending.

#### Web Resources

eMERGE: https://emerge.mc.vanderbilt.edu/

UKBB: https://www.ukbiobank.ac.uk/

BioMe: https://icahn.mssm.edu/research/ipm/programs/biome-biobank

PLINK: https://www.cog-genomics.org/plink2

PYTHON: https://www.python.org/

R: https://www.r-project.org/

KING: http://people.virginia.edu/~wc9c/KING/

FlashPCA: https://github.com/gabraham/flashpca

Michigan Imputation Server: https://imputationserver.sph.umich.edu

Human Reference Consortium: http://www.haplotype-reference-consortium.org

TOPMed Imputation Server: https://imputation.biodatacatalyst.nhlbi.nih.gov/

LDPred: https://github.com/bvilhjal/ldpred

GPS catalogue: https://www.pgscatalog.org/

## ACKNOWLEDGEMENTS

This work was funded by the National Human Genome Research Institute (NHGRI) Electronic Medical Records and Genomics-IV (eMERGE-IV grants 2U01HG008680-05, 1U01HG011167-01, 1U01HG011176-01). Additional sources of funding included UG3DK114926 (KK), RC2DK116690 (KK), R01LM013061 (CW, KK), K25DK128563 (AK) and UL1TR001873 (AK, KK), R01HL151855 (JBM), UM1DK078616 (JBM). The REGARDS (R01HL136666), HyperGEN (R01HL055673), GenHAT (R01HL123782), and WPC (R01HL092173, K24HL133373) studies were all supported by the National Heart, Lung, and Blood Institute (NHLBI). Parts of this study have been conducted using the UKBB Resource under UKBB project ID number 41849. The content is solely the responsibility of the authors and does not necessarily represent the official views of the National Institutes of Health.

## Supplementary Information

**Supplemental Figure S1:**
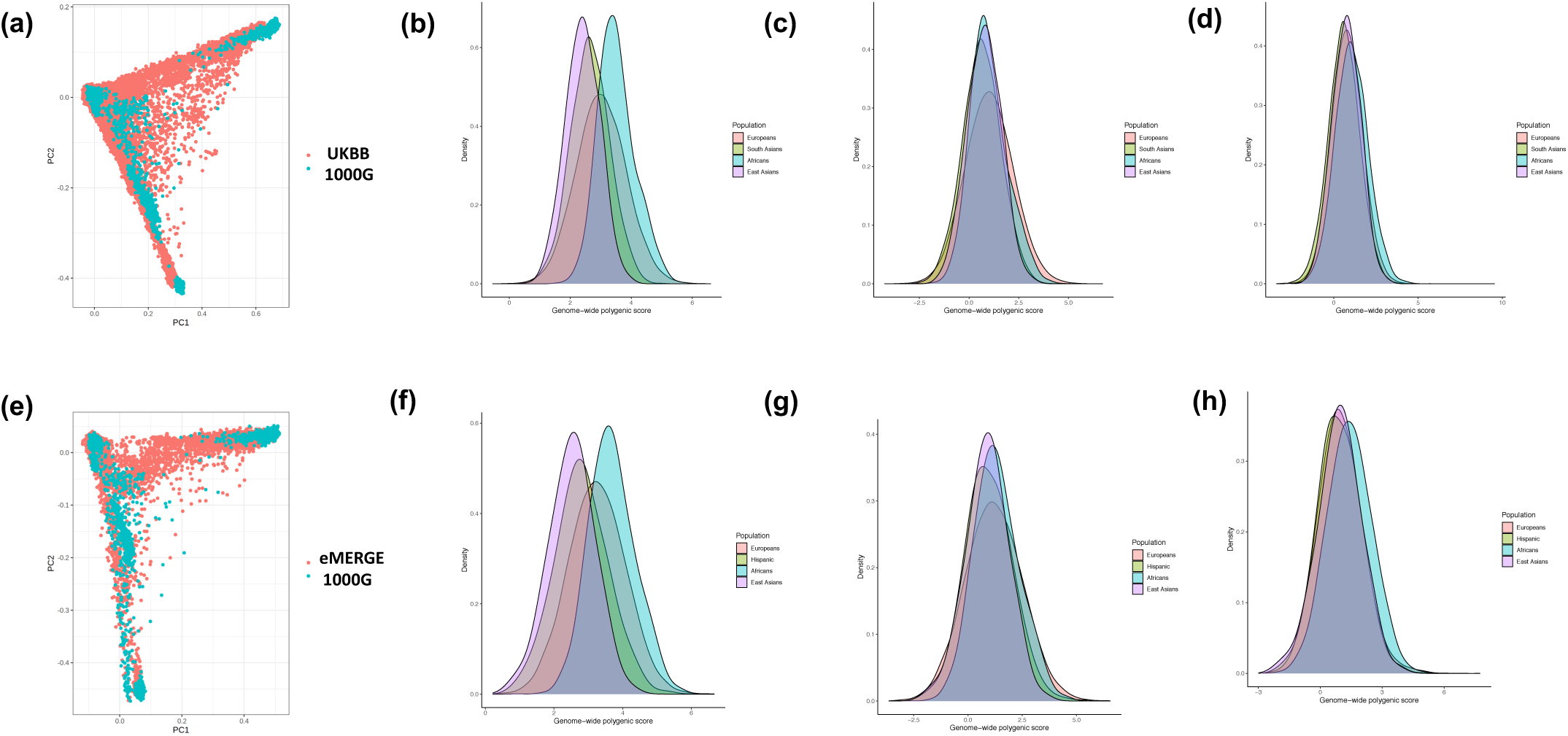
Risk score distributions eMERGE-III and UKBB datasets: **(a)** ancestry PCA projection of UKBB participants on all 1000G samples; **(b)** the distribution of raw polygenic score without *APOL1* in UKBB by ancestry; **(c)** the distribution of ancestry-adjusted polygenic score (method 1: mean-adjusted) in UKBB by ancestry; **(d)** the distribution of ancestry-adjusted polygenic score (method 2: mean and variance-adjusted) in UKBB by ancestry. Panels **(e), (f), (g)** and **(h)** show the same analyses for the eMERGE-III dataset, respectively.

**Supplemental Figure S2.**
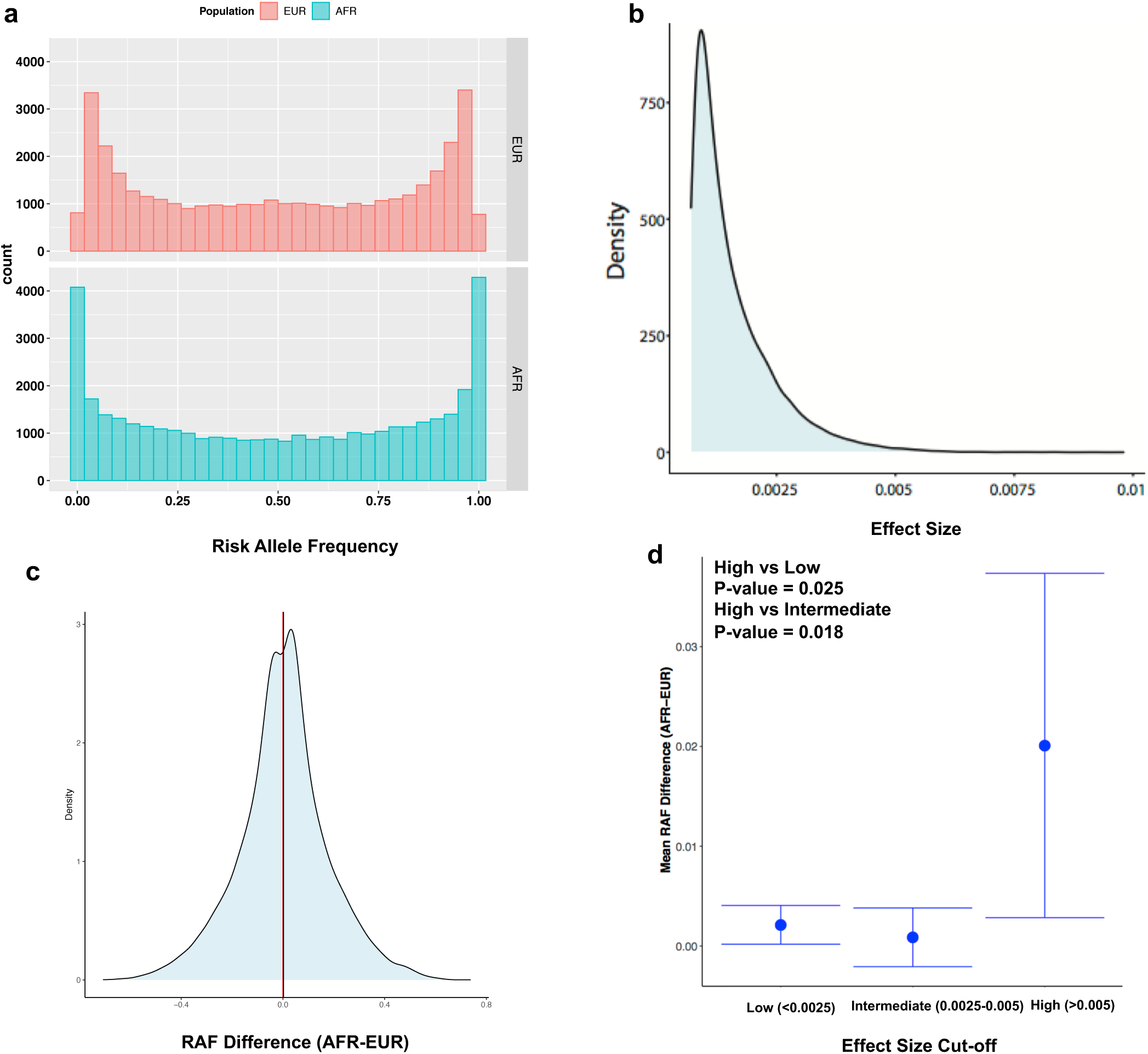
Distribution of risk allele frequencies (RAF) and their effect sizes for the variants included in the GPS. **(a)** comparison of RAF distributions for the risk variants included in the CKD GPS demonstrates higher frequency of rare (RAF<0.01) and common (RAF>0.99) risk alleles in African compared to European genomes (based on 1000G reference populations); this may be explained by the exclusion of variants with MAF<0.01 in European discovery GWAS; **(b)** highly skewed effect size (weight) distribution for the variants included in the GPS for CKD; **(c)** Distribution of RAF difference (AFR-EUR) demonstrating higher average frequency of risk alleles in African genomes (mean RAF difference = 0.002) and a slight rightward shift of the RAF difference distribution from the expected mean of 0; **(d)** Mean RAF difference (AFR-EUR) as a function of effect size binned into three categories (high, intermediate, and low) based on the observed distribution of effects sizes in panel b, demonstrating that the risk alleles with larger effect size have higher average frequency in African compared to European genomes. EUR: European and AFR: African. The bars represent 95% confidence intervals around the mean RAF difference estimate for each bin.

**Supplemental Figure S3:**
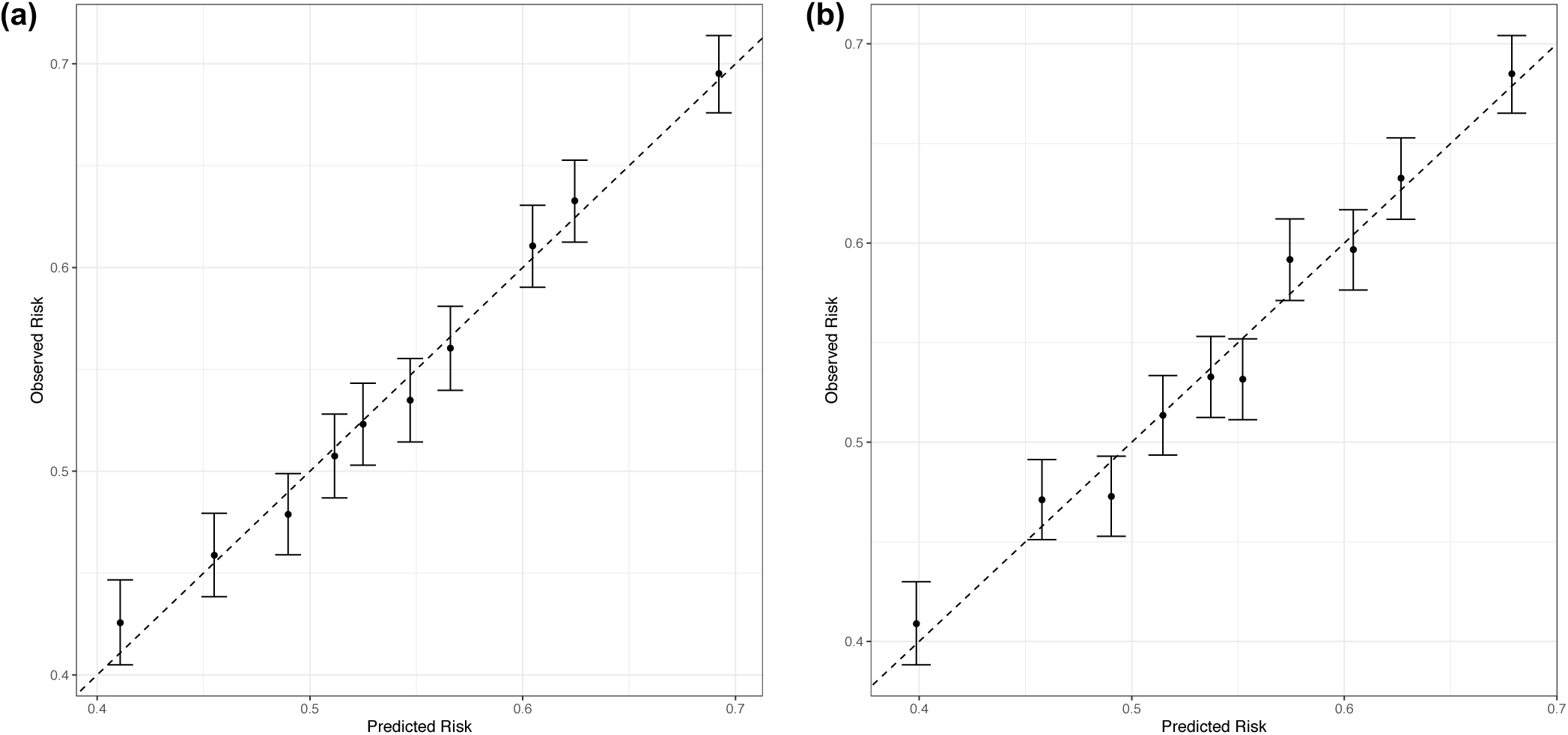
Final GPS calibration analysis in eMERGE-III cohorts combined: predicted risk (X-axis) as a function of the observed risk (Y-axis) in the multiethnic eMERGE-III dataset after ancestry adjustment with **(a)** method 1 and **(b)** method 2.

**Supplemental Figure S4:**
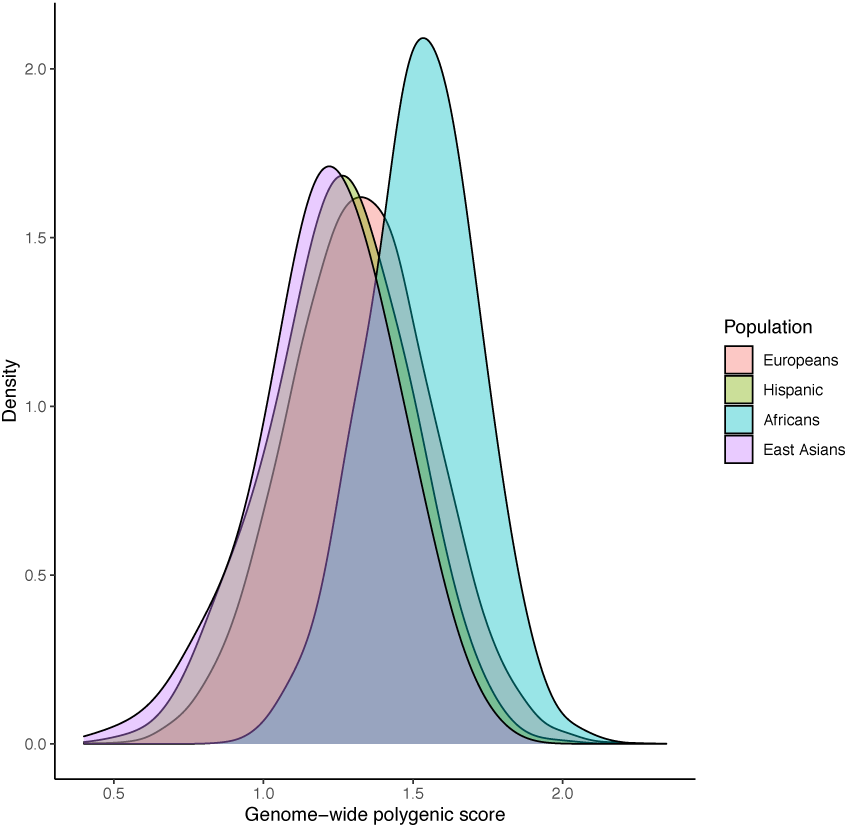
Distributions of the raw (non-standardized) genome-wide polygenic score (GPS) by Yu et al. in the eMERGE-III dataset by ancestry.

**Supplementary Table 1:**
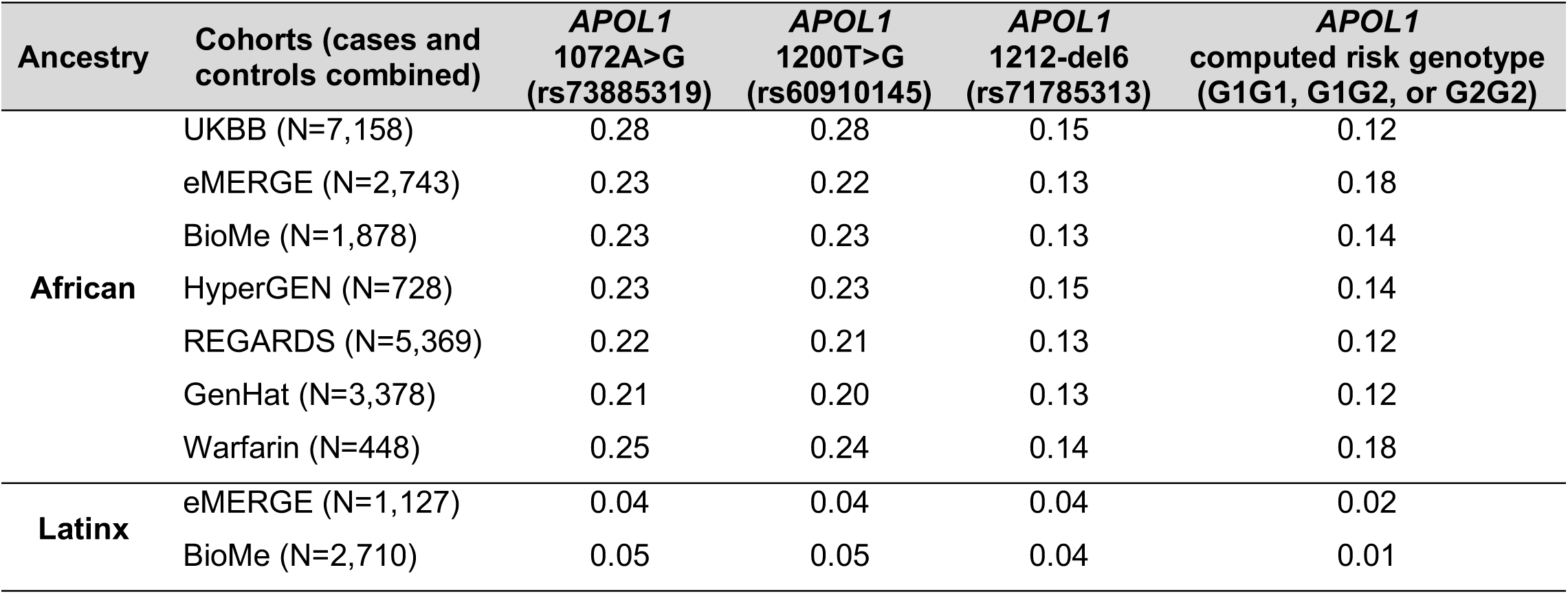
Overall frequencies of *APOL1* G1 and G2 risk alleles and *APOL1* risk genotypes in African American and Latinx cohorts included in the study.

| Ancestry | Cohorts (cases and controls combined) | *APOL1*<br>1072A>G<br>(rs73885319) | *APOL1*<br>1200T>G<br>(rs60910145) | *APOL1*<br>1212-del6<br>(rs71785313) | *APOL1*<br>computed risk genotype<br>(G1G1, G1G2, or G2G2) |
| --- | --- | --- | --- | --- | --- |
| **African** | UKBB (N=7,158) | 0.28 | 0.28 | 0.15 | 0.12 |
|  | eMERGE (N=2,743) | 0.23 | 0.22 | 0.13 | 0.18 |
|  | BioMe (N=1,878) | 0.23 | 0.23 | 0.13 | 0.14 |
|  | HyperGEN (N=728) | 0.23 | 0.23 | 0.15 | 0.14 |
|  | REGARDS (N=5,369) | 0.22 | 0.21 | 0.13 | 0.12 |
|  | GenHat (N=3,378) | 0.21 | 0.20 | 0.13 | 0.12 |
|  | Warfarin (N=448) | 0.25 | 0.24 | 0.14 | 0.18 |
| **Latinx** | eMERGE (N=1,127) | 0.04 | 0.04 | 0.04 | 0.02 |
|  | BioMe (N=2,710) | 0.05 | 0.05 | 0.04 | 0.01 |

**Supplementary Table 2:** Association of candidate polygenic scores with CKD in the first UKBB optimization dataset (70% of UKBB Europeans). Odds ratio (OR) per standard deviation (SD) of each risk score, and area under the receiver-operator curve (AUC) were calculated in the UKBB optimization dataset of 279,819 Europeans with adjustment for age, sex, diabetes, first four principal components of ancestry and genotyping batch; AUC crude and variance explained are calculated for the risk score component alone without any covariates; r^2^: linkage disequilibrium pruning threshold; ρ-tuning parameter to model the proportion of variants assumed to be causal; the best performing score is highlighted in red. Variance explained is estimated as a Nagelkerke pseudo R^2^ and refers to the variance in case-control status.

| Method | Tuning parameter | N variants | OR* per SD of GPS | P-value | AUC (Adjusted*) | AUC (Crude) | Variance Explained |
| --- | --- | --- | --- | --- | --- | --- | --- |
| P+T | P=1.0E-01 | 958,747 | 1.60 | P<1E-300 | 0.7244 | 0.6115 | 0.0301 |
| P+T | P=1.0E-02 | 280,537 | 1.60 | P<1E-300 | 0.7246 | 0.6115 | 0.0303 |
| P+T | P=1.0E-03 | 125,944 | 1.57 | P<1E-300 | 0.7224 | 0.6066 | 0.0276 |
| P+T | P=1.0E-04 | 73,199 | 1.56 | P<1E-300 | 0.7217 | 0.6047 | 0.0268 |
| P+T | P=1.0E-05 | 47,967 | 1.53 | P<1E-300 | 0.7199 | 0.6003 | 0.0248 |
| P+T | P=1.0E-06 | 33,414 | 1.52 | P<1E-300 | 0.7194 | 0.599 | 0.0241 |
| P+T | P=1.0E-07 | 23,534 | 1.52 | P<1E-300 | 0.7192 | 0.5986 | 0.0241 |
| P+T | P=1.0E-08 | 17,160 | 1.51 | P<1E-300 | 0.7181 | 0.5959 | 0.0229 |
| **P+T** | **P=3.0E-02** | **471,316** | **1.60** | **P<1E-300** | **0.7246** | **0.6118** | **0.0304** |
| P+T | P=3.0E-03 | 176,651 | 1.58 | P<1E-300 | 0.7236 | 0.6095 | 0.029 |
| P+T | P=3.0E-04 | 92,957 | 1.56 | P<1E-300 | 0.7221 | 0.6058 | 0.0273 |
| P+T | P=3.0E-05 | 58,204 | 1.54 | P<1E-300 | 0.7206 | 0.6021 | 0.0256 |
| LDPred | $\rho$=1.0E+00 | 5,440,627 | 1.58 | P<1E-300 | 0.7233 | 0.6093 | 0.0288 |
| LDPred | $\rho$=1.0E-01 | 5,440,627 | 1.15 | P=2.51E-97 | 0.6997 | 0.5346 | 0.0029 |
| LDPred | $\rho$=1.0E-02 | 5,440,627 | 1.12 | P=3.20E-60 | 0.6985 | 0.5273 | 0.0018 |
| LDPred | $\rho$=1.0E-03 | 5,440,627 | 1.11 | P=2.58E-56 | 0.6984 | 0.5266 | 0.0016 |
| LDPred | $\rho$=3.0E-01 | 5,440,627 | 1.51 | P<1E-300 | 0.7183 | 0.5975 | 0.0232 |
| LDPred | $\rho$=3.0E-02 | 5,440,627 | 1.08 | P=4.31E-30 | 0.6976 | 0.5187 | 9.00E-04 |
| LDPred | $\rho$=3.0E-03 | 5,440,627 | 1.12 | P=9.13E-58 | 0.6985 | 0.5263 | 0.0017 |

\* Adjusted for age, sex, diabetes, the first four principal components of ancestry and genotyping batch.

**Supplementary Table 3:** Mutually-adjusted effects for *APOL1* risk genotype and the best GPS from the first optimization cohort when tested in the second optimization cohort of African ancestry.

| Model * | Effect ($\beta$) | 95% CI | P-value |
| --- | --- | --- | --- |
| Standardized GPS | 0.15 | 0.09-0.22 | P=1.0E-04 |
| *APOL1* risk genotype | 0.17 | 0.01-0.32 | P=4.0E-02 |
| GPS and *APOL1* risk genotype interaction | 0.09 | -0.07-0.25 | P=0.29 (NS) |

\* adjusted for age, sex, diabetes, four principal components of ancestry, and genotyping batch.

**Supplemental Table 4:** GPS performance meta-analysis for testing cohorts of European ancestry (23,364 cases and 117,883 controls in total)

| Cohort | Case/control | OR per SD (95% CI), P-value | AUC (Crude) | PRS Threshold | Odds ratio (95% CI), P value |
| --- | --- | --- | --- | --- | --- |
| UKBB | 11,922/108,002 | 1.60 (1.58-1.62), P<1.0E-300 | 0.72 (0.61) | Top 20% vs. other 80% | 2.23 (2.18-2.28), P=4.33E-254 |
|  |  |  |  | Top 10% vs. other 90% | 2.48 (2.42-2.54), P=1.42E-206 |
|  |  |  |  | Top 5% vs. other 95% | 2.72 (2.64-2.80), P=8.46E-144 |
|  |  |  |  | **Top 2% vs. other 98%** | **3.16 (3.04-3.28), P=3.66E-86** |
|  |  |  |  | Top 1% vs. other 99% | 3.67 (3.51-3.83), P=1.29E-58 |
| eMERGE | 10,572/8,030 | 1.38 (1.35-1.40), P=1.58E-83 | 0.77 (0.60) | Top 20% vs. other 80% | 1.84 (1.69-2.00), P=1.54E-45 |
|  |  |  |  | Top 10% vs. other 90% | 1.90 (1.70-2.14), P=5.87E-28 |
|  |  |  |  | Top 5% vs. other 95% | 2.15 (1.83-2.54), P=7.49E-20 |
|  |  |  |  | **Top 2% vs. other 98%** | **2.65 (2.02-3.48), P=1.35E-12** |
|  |  |  |  | Top 1% vs. other 99% | 3.22 (2.17-4.78), P=6.80E-09 |
| BioME | 870/1,851 | 1.58 (1.46-1.70), P=2.50E-14 | 0.91 (0.65) | Top 20% vs. other 80% | 2.42 (2.13-2.71), P=1.39E-09 |
|  |  |  |  | Top 10% vs. other 90% | 2.31 (1.94-2.68), P=7.88E-06 |
|  |  |  |  | Top 5% vs. other 95% | 1.99 (1.50-2.48), P=5.57E-03 |
|  |  |  |  | **Top 2% vs. other 98%** | **3.39 (2.60-4.18), P=2.54E-03** |
|  |  |  |  | Top 1% vs. other 99% | 5.87 (4.74-7.00), P=2.01E-03 |
| Meta | 23,364/117,883 | 1.49 (1.47-1.50), P<1.0E-300 | 0.75 (0.61) | Top 20% vs. other 80% | 2.14 (2.10-2.18), P=1.04E-300 |
|  |  |  |  | Top 10% vs. other 90% | 2.35 (2.30-2.40), P=6.09E-234 |
|  |  |  |  | Top 5% vs. other 95% | 2.59 (2.52-2.66), P=3.83E-161 |
|  |  |  |  | **Top 2% vs. other 98%** | **3.09 (2.98-3.19), P=3.22E-98** |
|  |  |  |  | Top 1% vs. other 99% | 3.63 (3.49-3.78), P=5.09E-68 |

OR: Odds ratio for the model adjusted for age, sex, diabetes, principal components of ancestry and genotyping array or clinical site; SD: standard deviation of the GPS distribution in controls; AUC: area under the receiver-operator curve for the model adjusted for age, sex, diabetes, principal components of ancestry and genotyping array or clinical site (crude: AUC for GPS alone without any covariates).

**Supplementary Table 5:**
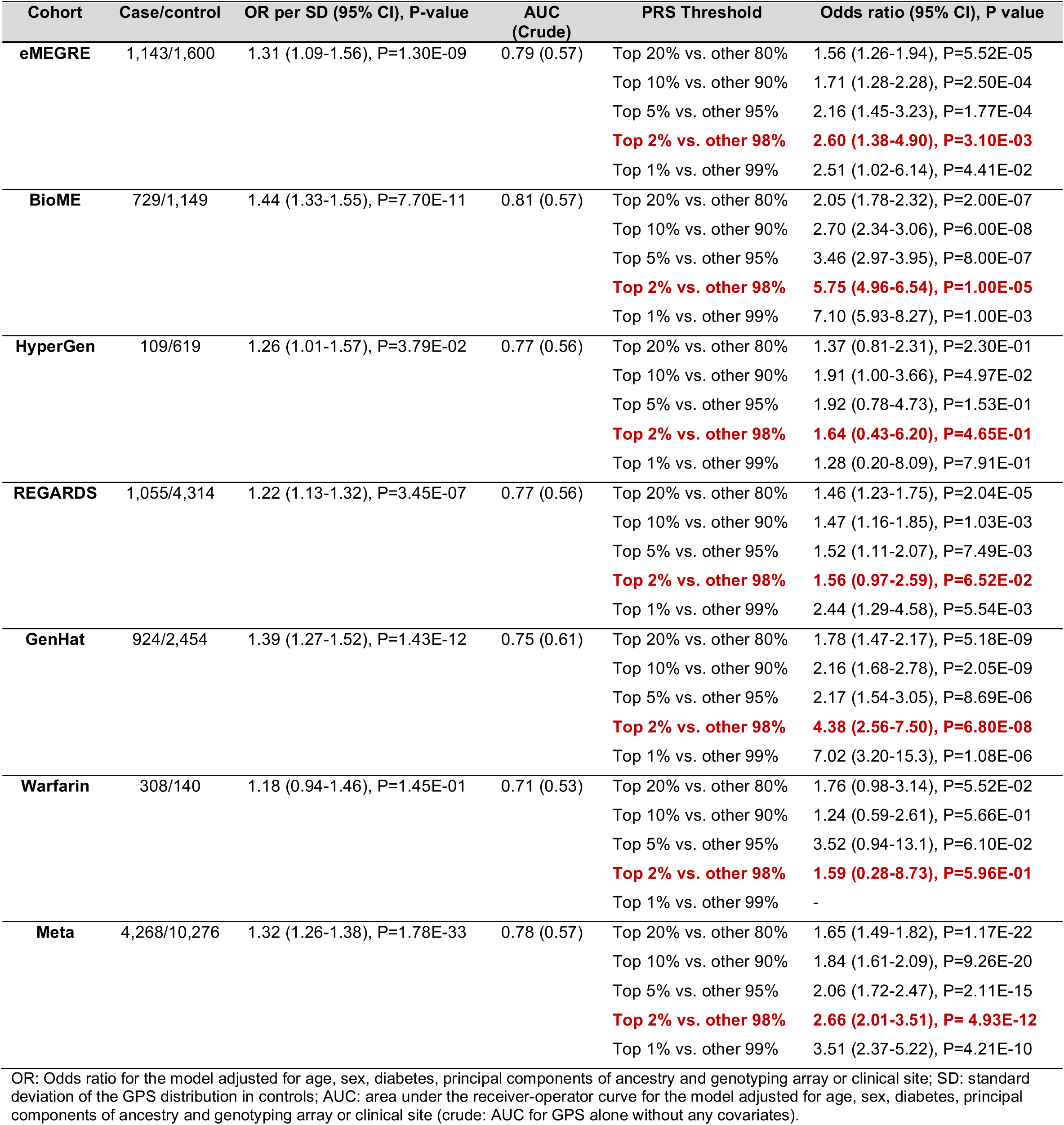
GPS performance meta-analysis for testing cohorts of African ancestry (4,268 cases and 10,276 controls in total).

| Cohort | Case/control | OR per SD (95% CI), P-value | AUC (Crude) | PRS Threshold | Odds ratio (95% CI), P value |
| --- | --- | --- | --- | --- | --- |
| **eMEGRE** | 1,143/1,600 | 1.31 (1.09-1.56), P=1.30E-09 | 0.79 (0.57) | Top 20% vs. other 80% | 1.56 (1.26-1.94), P=5.52E-05 |
|  |  |  |  | Top 10% vs. other 90% | 1.71 (1.28-2.28), P=2.50E-04 |
|  |  |  |  | Top 5% vs. other 95% | 2.16 (1.45-3.23), P=1.77E-04 |
|  |  |  |  | **Top 2% vs. other 98%** | **2.60 (1.38-4.90), P=3.10E-03** |
|  |  |  |  | Top 1% vs. other 99% | 2.51 (1.02-6.14), P=4.41E-02 |
| **BioME** | 729/1,149 | 1.44 (1.33-1.55), P=7.70E-11 | 0.81 (0.57) | Top 20% vs. other 80% | 2.05 (1.78-2.32), P=2.00E-07 |
|  |  |  |  | Top 10% vs. other 90% | 2.70 (2.34-3.06), P=6.00E-08 |
|  |  |  |  | Top 5% vs. other 95% | 3.46 (2.97-3.95), P=8.00E-07 |
|  |  |  |  | **Top 2% vs. other 98%** | **5.75 (4.96-6.54), P=1.00E-05** |
|  |  |  |  | Top 1% vs. other 99% | 7.10 (5.93-8.27), P=1.00E-03 |
| **HyperGen** | 109/619 | 1.26 (1.01-1.57), P=3.79E-02 | 0.77 (0.56) | Top 20% vs. other 80% | 1.37 (0.81-2.31), P=2.30E-01 |
|  |  |  |  | Top 10% vs. other 90% | 1.91 (1.00-3.66), P=4.97E-02 |
|  |  |  |  | Top 5% vs. other 95% | 1.92 (0.78-4.73), P=1.53E-01 |
|  |  |  |  | **Top 2% vs. other 98%** | **1.64 (0.43-6.20), P=4.65E-01** |
|  |  |  |  | Top 1% vs. other 99% | 1.28 (0.20-8.09), P=7.91E-01 |
| **REGARDS** | 1,055/4,314 | 1.22 (1.13-1.32), P=3.45E-07 | 0.77 (0.56) | Top 20% vs. other 80% | 1.46 (1.23-1.75), P=2.04E-05 |
|  |  |  |  | Top 10% vs. other 90% | 1.47 (1.16-1.85), P=1.03E-03 |
|  |  |  |  | Top 5% vs. other 95% | 1.52 (1.11-2.07), P=7.49E-03 |
|  |  |  |  | **Top 2% vs. other 98%** | **1.56 (0.97-2.59), P=6.52E-02** |
|  |  |  |  | Top 1% vs. other 99% | 2.44 (1.29-4.58), P=5.54E-03 |
| **GenHat** | 924/2,454 | 1.39 (1.27-1.52), P=1.43E-12 | 0.75 (0.61) | Top 20% vs. other 80% | 1.78 (1.47-2.17), P=5.18E-09 |
|  |  |  |  | Top 10% vs. other 90% | 2.16 (1.68-2.78), P=2.05E-09 |
|  |  |  |  | Top 5% vs. other 95% | 2.17 (1.54-3.05), P=8.69E-06 |
|  |  |  |  | **Top 2% vs. other 98%** | **4.38 (2.56-7.50), P=6.80E-08** |
|  |  |  |  | Top 1% vs. other 99% | 7.02 (3.20-15.3), P=1.08E-06 |
| **Warfarin** | 308/140 | 1.18 (0.94-1.46), P=1.45E-01 | 0.71 (0.53) | Top 20% vs. other 80% | 1.76 (0.98-3.14), P=5.52E-02 |
|  |  |  |  | Top 10% vs. other 90% | 1.24 (0.59-2.61), P=5.66E-01 |
|  |  |  |  | Top 5% vs. other 95% | 3.52 (0.94-13.1), P=6.10E-02 |
|  |  |  |  | **Top 2% vs. other 98%** | **1.59 (0.28-8.73), P=5.96E-01** |
|  |  |  |  | Top 1% vs. other 99% | - |
| **Meta** | 4,268/10,276 | 1.32 (1.26-1.38), P=1.78E-33 | 0.78 (0.57) | Top 20% vs. other 80% | 1.65 (1.49-1.82), P=1.17E-22 |
|  |  |  |  | Top 10% vs. other 90% | 1.84 (1.61-2.09), P=9.26E-20 |
|  |  |  |  | Top 5% vs. other 95% | 2.06 (1.72-2.47), P=2.11E-15 |
|  |  |  |  | **Top 2% vs. other 98%** | **2.66 (2.01-3.51), P=4.93E-12** |
|  |  |  |  | Top 1% vs. other 99% | 3.51 (2.37-5.22), P=4.21E-10 |

OR: Odds ratio for the model adjusted for age, sex, diabetes, principal components of ancestry and genotyping array or clinical site; SD: standard deviation of the GPS distribution in controls; AUC: area under the receiver-operator curve for the model adjusted for age, sex, diabetes, principal components of ancestry and genotyping array or clinical site (crude: AUC for GPS alone without any covariates).

**Supplementary Table 6:** GPS performance meta-analysis for testing cohorts of Latinx/Hispanic ancestry (1,492 cases and 2,984 controls in total).

| Cohort | Case/control | OR per SD (95% CI), P-value | AUC (Crude) | PRS Threshold | Odds ratio (95% CI), P value |
| --- | --- | --- | --- | --- | --- |
| **eMEGRE** | 488/639 | 1.39 (1.22-1.56), P=1.04E-04 | 0.87 (0.57) | Top 20% vs. other 80% | 2.56 (1.71-3.81), P=4.06E-06 |
|  |  |  |  | Top 10% vs. other 90% | 2.42 (1.43-4.07), P=8.69E-04 |
|  |  |  |  | Top 5% vs. other 95% | 2.83 (1.36-5.89), P=5.25E-03 |
|  |  |  |  | **Top 2% vs. other 98%** | **3.82 (1.15-12.62), P=2.83E-02** |
|  |  |  |  | Top 1% vs. other 99% | 7.61 (1.22-47.37), P=2.96E-02 |
| **BioME** | 1,004/2,345 | 1.47 (1.35-1.59), P=8.0E-10 | 0.89 (0.63) | Top 20% vs. other 80% | 1.74 (1.47-2.01), P=5.34E-05 |
|  |  |  |  | Top 10% vs. other 90% | 2.26 (1.90-2.62), P=7.77E-06 |
|  |  |  |  | Top 5% vs. other 95% | 2.77 (2.27-3.27), P=6.27E-05 |
|  |  |  |  | **Top 2% vs. other 98%** | **4.48 (3.69-5.27), P=2.10E-04** |
|  |  |  |  | Top 1% vs. other 99% | 6.30 (5.13-7.47), P=2.07E-02 |
| **Meta** | 1,492/2,984 | 1.42 (1.33-1.51), P=4.10E-14 | 0.88 (0.61) | Top 20% vs. other 80% | 1.96 (1.57-2.46), P=3.40E-09 |
|  |  |  |  | Top 10% vs. other 90% | 2.31 (1.72-3.11), P=3.01E-08 |
|  |  |  |  | Top 5% vs. other 95% | 2.79 (1.85-4.22), P=1.13E-06 |
|  |  |  |  | **Top 2% vs. other 98%** | **4.27 (2.21-8.25), P=1.59E-05** |
|  |  |  |  | Top 1% vs. other 99% | 6.66 (2.48-17.8), P=1.63E-04 |

OR: Odds ratio for the model adjusted for age, sex, diabetes, principal components of ancestry and genotyping array or clinical site; SD: standard deviation of the GPS distribution in controls; AUC: area under the receiver-operator curve for the model adjusted for age, sex, diabetes, principal components of ancestry and genotyping array or clinical site (crude: AUC for GPS alone without any covariates).

**Supplementary Table 7:** GPS performance meta-analysis for testing cohorts of Asian continental ancestry (1,030 cases and 9,896 controls in total).

| Cohort | Case/Control | OR per SD (95% CI), P-value | AUC (Crude) | PRS Threshold | Odds ratio (95% CI), P value |
| --- | --- | --- | --- | --- | --- |
| **eMERGE (E.Asian)** | 98/105 | 1.90 (1.43-2.37), P=7.00E-03 | 0.92 (0.59) | Top 20% vs. other 80% | 3.86 (1.23-12.07), P=2.05E-02 |
|  |  |  |  | Top 10% vs. other 90% | 5.93 (1.44-24.32), P=1.33E-02 |
|  |  |  |  | Top 5% vs. other 95% | 5.58 (3.75-7.41), P=6.57E-02 |
|  |  |  |  | **Top 2% vs. other 98%** | **10.6 (0.58-192.8), P=1.10E-01** |
|  |  |  |  | Top 1% vs. other 99% | 2.86 (0.04-205.1), P=6.31E-01 |
| **UKBB (E.Asian)** | 74/1,740 | 1.46 (1.17-1.75), P=1.00E-02 | 0.86 (0.58) | Top 20% vs. other 80% | 1.27 (0.61-1.93), P=4.84E-01 |
|  |  |  |  | Top 10% vs. other 90% | 1.41 (0.60-2.22), P=4.07E-01 |
|  |  |  |  | Top 5% vs. other 95% | 2.21 (1.23-3.19), P=1.15E-01 |
|  |  |  |  | **Top 2% vs. other 98%** | **3.32 (1.85-4.79), P=1.10E-01** |
|  |  |  |  | Top 1% vs. other 99% | 4.90 (3.03-6.77), P=9.42E-02 |
| **UKBB (SW Asian)** | 797/7,698 | 1.58 (1.50-1.66), P=3.50E-27 | 0.75 (0.60) | Top 20% vs. other 80% | 2.14 (1.96-2.32), P=4.01E-16 |
|  |  |  |  | Top 10% vs. other 90% | 2.37 (2.14-2.60), P=1.78E-13 |
|  |  |  |  | Top 5% vs. other 95% | 2.66 (2.36-2.96), P=1.76E-10 |
|  |  |  |  | **Top 2% vs. other 98%** | **2.70 (2.24-3.16), P=2.06E-05** |
|  |  |  |  | Top 1% vs. other 99% | 3.49 (2.87-4.11), P=8.25E-05 |
| **BioMe (E. Asian)** | 61/353 | 1.89 (1.43-2.35), P=6.89E-03 | 0.89 (0.63) | Top 20% vs. other 80% | 2.61 (1.66-3.56), P=4.83E-02 |
|  |  |  |  | Top 10% vs. other 90% | 4.57 (3.41-5.73), P=1.05E-01 |
|  |  |  |  | Top 5% vs. other 95% | 3.13 (1.11-5.15), P=2.65E-01 |
|  |  |  |  | **Top 2% vs. other 98%** | **10.1 (7.71-12.5), P=5.78E-02** |
|  |  |  |  | Top 1% vs. other 99% | 20.9 (18.3-23.5), P=2.26E-02 |
| **Meta** | 1,030/9,896 | 1.59 (1.52-1.67), P=1.30E-30 | 0.90 (0.60) | Top 20% vs. other 80% | 2.03 (1.71-2.41), P=3.76E-16 |
|  |  |  |  | Top 10% vs. other 90% | 2.38 (1.93-2.96), P=1.69E-15 |
|  |  |  |  | Top 5% vs. other 95% | 2.67 (2.01-3.54), P=6.42E-12 |
|  |  |  |  | **Top 2% vs. other 98%** | **2.95 (1.93-4.50), P=5.91E-07** |
|  |  |  |  | Top 1% vs. other 99% | 3.91 (2.22-6.91), P=2.66E-06 |

OR: Odds ratio for the model adjusted for age, sex, diabetes, principal components of ancestry and genotyping array or clinical site; SD: standard deviation of the GPS distribution in controls; AUC: area under the receiver-operator curve for the model adjusted for age, sex, diabetes, principal components of ancestry and genotyping array or clinical site (crude: AUC for GPS alone without any covariates).

**Supplementary Table 8:**
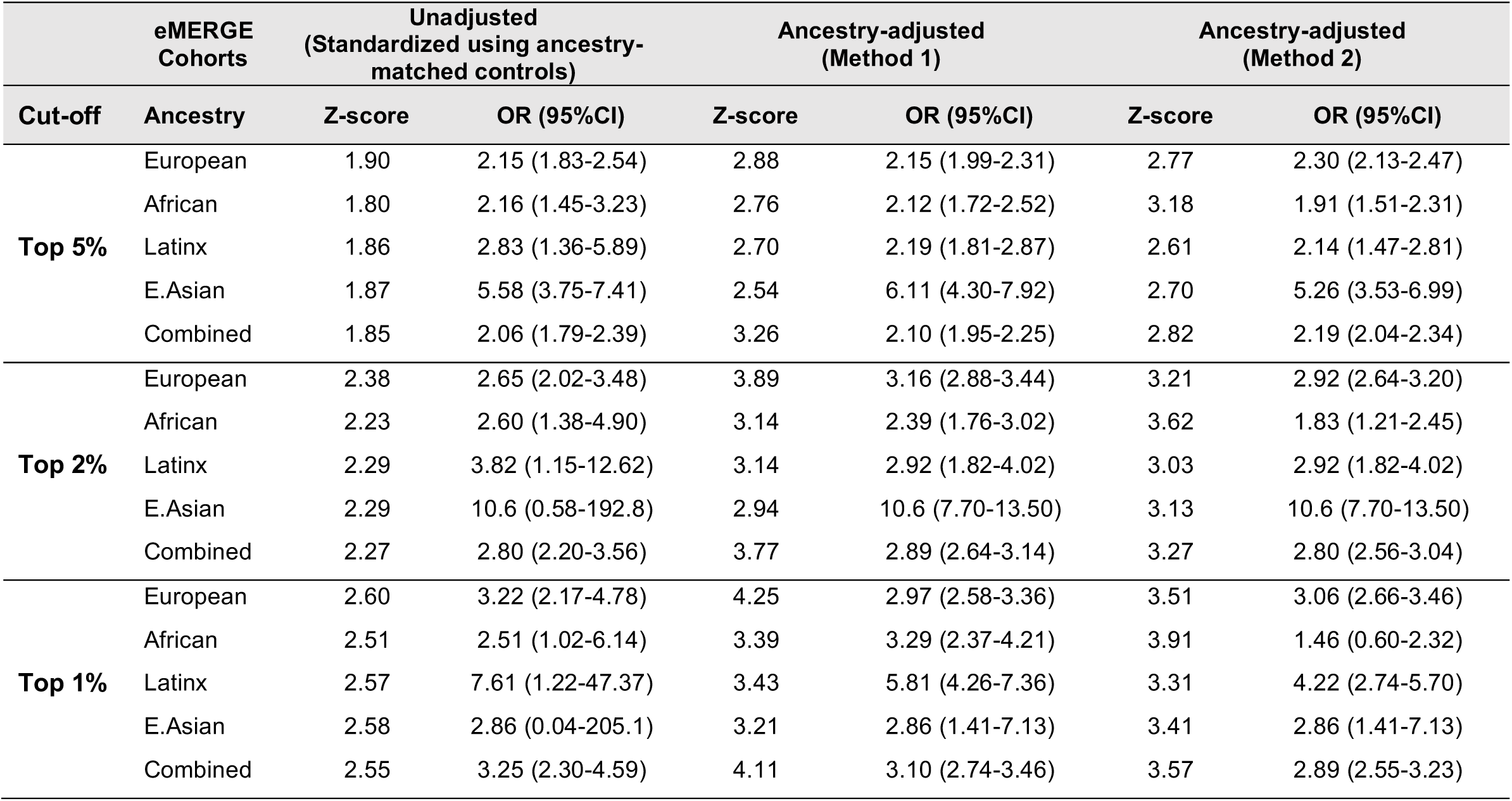
Comparison of the changes in GPS effects for the top 1%, 2%, and 5% tail cut-offs using different ancestry adjustment methods in the eMERGE-III dataset.

**Supplementary Table 9.** Comparison of the design, optimization, and testing strategies for the polygenic risk score published by Yu et al. with the score from the present study.

|  | Yu et al. (*JASN* 2021) | Khan et al. (*present study*) |
| --- | --- | --- |
| **Design** |  |  |
| GWAS summary statistics | CKDGen without ARIC + 90% of UKBB | CKDGen meta-analysis GWAS for eGFR |
| GWAS phenotype | Estimated GFR | Estimated GFR |
| GWAS sample size | N=1,159,871 | N=765,348 |
| GWAS ancestral composition | 82% European<br>16% Asian<br>1.5% African<br><1% Hispanic | 75% European<br>23% Asian<br>2% African<br><1% Hispanic |
| External LD reference | N=608 (1000G multiethnic subset) | N=2,504 (1000G all populations) |
| APOL1 risk genotype | Not included | Included under a recessive model |
| Covariates | Age, sex, diabetes, PCs of ancestry | Age, sex, diabetes, PCs of ancestry |
| **Optimization** |  |  |
| Best performing model | LDpred (1.5M variants) | P+T (471K variants) |
| Optimization datasets | 10% of UKBB (N=45,158) | 70% UKBB European ancestry (N=279,819) AND<br>100% UKBB African ancestry (N=7,158) |
| **Testing** |  |  |
| Overall Strategy | ARIC prospective cohort (total N=11,737) | 15 Case-Control Cohorts (total N=171,213) |
| Outcome definition | Incident CKD (eGFR <60 mL/min/1.73 m<sup>2</sup> plus ≥30% eGFR decline during follow-up). | CKD stage 3 or above (eGFR <60 mL/min/1.73) including ESRD (chronic dialysis, transplant) |
| European ancestry | ARIC Europeans (N=8,866) | 3 European ancestry cohorts (N=141,247) |
| African ancestry | ARIC African Americans (N=2,871) | 6 African ancestry cohorts (N=14,544) |
| Hispanic/Latinx ancestry | NA | 2 Latinx cohorts (N=4,476) |
| Asian ancestry | NA | 4 Asian cohorts (N=10,926) |

**Supplementary Table 10.** Comparison of the performance characteristics of the polygenic score published by Yu et al. with the score from the present study across the ancestry-defined eMERGE-III case-control cohorts. Adjusted AUC refers to the area under the receiver operating curve for the full model (PRS, age, sex, diabetes, and ancestry PCs). Crude AUC (in parenthesis) refers to the area under the receiver operating curve for the PRS predictor alone. Variance explained was estimated as a Nagelkerke pseudo-R^2^ and refers to the variance in case-control status explained by the PRS predictor alone. Performance differences in the African-ancestry testing cohort highlighted in red.

| Performance Metrics | Yu et al. (*JASN* 2021) | Khan et al. (*present study*) |
| --- | --- | --- |
| **Adjusted AUC (Crude AUC), Pseudo $R^2$** |  |  |
| European (N case/control = 10,572/8,030) | AUC 0.78 (0.61), $R^2$ 0.035 | AUC 0.77 (0.60), $R^2$ 0.022 |
| African (N case/control = 1,143/1,600) | AUC 0.78 (0.55), $R^2$ 0.010 | AUC 0.78 (0.57), $R^2$ 0.014 |
| Latinx (N case/control = 488/639) | AUC 0.87 (0.58), $R^2$ 0.012 | AUC 0.87 (0.57), $R^2$ 0.011 |
| Asian (N case/control = 98/105) | AUC 0.91 (0.58), $R^2$ 0.010 | AUC 0.92 (0.59), $R^2$ 0.026 |
| **OR per SD (95% CI), P-value** |  |  |
| European (N case/control = 10,572/8,030) | 1.51 (1.46-1.56), P=6.25E-129 | 1.38 (1.35-1.40), P=1.58E-83 |
| African (N case/control = 1,143/1,600) | 1.28 (1.17-1.41), P=1.07E-07 | 1.31 (1.09-1.56), P=1.30E-09 |
| Latinx (N case/control = 488/639) | 1.45 (1.21-1.73), P=6.77E-05 | 1.39 (1.22-1.56), P=1.04E-04 |
| Asian (N case/control = 98/105) | 1.53 (0.94-2.49), P=8.60E-02 | 1.90 (1.43-2.37), P=7.00E-03 |
| **OR for top 2% (95% CI), P-value** |  |  |
| European (N case/control = 10,572/8,030) | 3.60 (2.70-4.79), P=1.25E-18 | 2.65 (2.02-3.48), P=1.35E-12 |
| African (N case/control = 1,143/1,600) | 1.92 (1.03-3.59), P=3.93E-02 | 2.60 (1.38-4.90), P=3.10E-03 |
| Latinx (N case/control = 488/639) | 1.35 (0.44-4.17), P=6.05E-01 | 3.82 (1.15-12.6), P=2.83E-02 |
| Asian (N case/control = 98/105) | 2.92 (0.07-118.0), P=5.70E-01 | 10.6 (0.58-192.8), P=1.10E-01 |

